# Personal HbA1c trajectories and incident diabetes: association, prediction and observability in the English Longitudinal Study of Ageing

**DOI:** 10.64898/2026.11.20.26368578

**Authors:** Diego Tapias, Cédric Donati, Susanne Völter-Mahlknecht

**Affiliations:** Department of Occupational, Social and Preventive Medicine, University Medical Center Göttingen, Georg-August-University Göttingen, Waldweg 37 A, 37073 Göttingen, Germany

**Keywords:** HbA1c, incident diabetes, personalised reference intervals, biological variation, longitudinal biomarkers, individual trajectories, Ornstein–Uhlenbeck process, risk prediction, ELSA

## Abstract

**Background:** Clinical diagnosis often relies on a snapshot: a single measurement interpreted against a population threshold. Repeated (longitudinal) measurements, however, may reveal an individual’s typical level and how it changes over time. Extracting this information is challenging when measurements are sparse and biological fluctuations may resemble sustained changes. Using data from the English Longitudinal Study of Ageing (ELSA), we investigated whether individual glycated haemoglobin (HbA1c) trajectories improved the prediction of incident diabetes beyond the most recent measurement, whether they were associated with incident diabetes, and how reliably they could be estimated.

**Methods:** In this secondary analysis of a prospective cohort study, we analysed venous HbA1c measured approximately every four years in ELSA, with a longer final interval of about seven years. A linear mixed model estimated each person’s usual HbA1c level and rate of change (personal slope) from their measurements before each diabetes-free health visit, with model parameters estimated in other participants (cross-fitting). We derived four longitudinal features: deviation from the person’s usual level, personal slope, change since the last measurement, and the forecast probability that the next HbA1c reaches the diagnostic threshold for diabetes. We evaluated the associations of these features with incident diabetes and their incremental predictive value beyond baseline HbA1c and clinical risk factors in internal, cross-fitted validation, compared predictive performance with a self-supervised Transformer, and modelled within-person variation as serially correlated drift (an Ornstein– Uhlenbeck process) to assess how reliably personal slopes can be recovered under different measurement schedules.

**Results:** The analysis included 5,828 diabetes-free baseline visits from 3,603 participants, of which 234 were followed by incident diabetes. Neither the engineered longitudinal features nor a Transformer that also used additional clinical variables improved discrimination beyond baseline HbA1c and clinical risk factors (difference in the area under the receiver operating characteristic curve, ΔAUC, *−*0.002 to +0.010; all 95% confidence intervals [CIs] including zero). The estimated personal slope was nevertheless associated with incident diabetes after adjustment for baseline HbA1c, age and other clinical factors (odds ratio [OR] per standard deviation [SD] 1.18, 95% CI 1.02–1.36, and 1.39, 1.11–1.74, among visits with at least two earlier HbA1c values), whereas deviation from the person’s usual level was not (OR 0.99, 95% CI 0.81–1.21). In the drift model, serially correlated fluctuations accounted for 77–81% of residual within-person variance, although the estimated temporal structure was sensitive to the treatment of the first HbA1c values at or above the diagnostic threshold. Under this model, persistent individual trends were difficult to distinguish from slowly varying fluctuations: with the intercept–slope correlation set to zero, estimated and underlying slopes correlated weakly for four-yearly measurements over up to 12 years (*r ≤* 0.26), and more frequent measurements improved this little.

**Conclusions:** Estimated personal HbA1c slopes carried adjusted prognostic information beyond baseline HbA1c and clinical factors, but longitudinal features, including those learned by a Transformer, did not clearly improve prediction. The drift model suggests that slowly varying within-person fluctuations may limit the reliable estimation of individual slopes from sparse measurements, and that longer observation histories are more informative than more frequent measurement. Whether personal laboratory histories improve prediction depends not only on their association with disease, but also on how reliably individual change can be recovered from the available measurements.

## 1 Introduction

Laboratory diagnosis often relies on comparing a single measurement with a population reference interval or a fixed diagnostic threshold (Clinical and Laboratory Standards Institute, 2010; World Health Organization, 2011). Yet many analytes vary far less within individuals than between them (Harris, 1974; Fraser, 2001; Ricós et al., 1999), and laboratory measurements are often repeated over years. Viewed longitudinally, these measurements may reveal what a single measurement cannot: an individual’s typical level and how it changes over time. A value considered normal for the population may therefore be unusual for a particular individual or reflect a sustained change from their personal baseline. Recent studies have demonstrated stable individual laboratory setpoints, their prognostic relevance, and the potential of Bayesian and machine-learning methods to construct personalised reference intervals (Foy et al., 2025; Zhang et al., 2026; Shah et al., 2026), building on earlier work in personal profiling (Chen et al., 2012; Price et al., 2017; Schüssler-Fiorenza Rose et al., 2019).

Three different claims are often merged under the label “personal reference”. The *deviation claim* is that a value unusually high relative to the person’s own baseline signals emerging disease. It is motivated by low indices of individuality (Harris, 1974; Fraser and Harris, 1989) and underlies personalised reference intervals (Coşkun et al., 2021) and the athlete biological passport, which flags values outside Bayesian person-specific limits (Sottas et al., 2011). The *trajectory claim* is that the direction and rate at which a person’s values are moving —their personal slope— carries risk information beyond the current level. The best-known example is prostate-specific antigen (PSA) velocity, proposed as a marker of prostate cancer (Carter et al., 1992); for glycaemia, fasting glucose and HbA1c rise steeply in the years before diagnosis of type 2 diabetes (Tabák et al., 2009; Heianza et al., 2012). Yet where the trajectory claim has been tested against the current level, the added value has often been small: a systematic review found little evidence that PSA velocity improves prediction beyond PSA itself (Vickers et al., 2009). The third use of history is *denoising*: earlier measurements improve estimation of the current underlying level (Irwig et al., 1991; Glasziou et al., 2008; Robinson, 1991). A feature built from earlier values can therefore appear to carry deviation or trajectory information merely because it refines the estimate of the current level, and an apparent change between two values partly reflects regression to the mean (Barnett et al., 2005). These three uses of history have different implications for monitoring, yet they are rarely separated empirically.

Distinguishing these effects also depends on how longitudinal measurements are collected. Electronic health records (EHRs) provide repeated observations, but tests are typically ordered for clinical reasons, making the timing and frequency of measurements informative. In one large hospital system, the timing of a test predicted survival better than its result for most laboratory tests (Agniel et al., 2018). Moreover, individuals who undergo repeated testing may differ systematically from those who do not, potentially distorting risk associations through informed presence (Goldstein et al., 2016; Hripcsak and Albers, 2013; Sisk et al., 2021). Population cohorts with scheduled measurements reduce the influence of clinically triggered testing (Steptoe et al., 2013), but introduce a different limitation: observations are typically sparse. Under such conditions, the reliability of estimated individual rates of change depends on the duration of follow-up and the number and precision of measurements (Willett, 1989; Raudenbush and Liu, 2001; Rast and Hofer, 2014; Brandmaier et al., 2018). How these constraints interact with biological fluctuations that persist over years remains poorly understood.

Even when individual trajectories can be estimated, their association with disease does not necessarily translate into improved prediction. A marker may be associated with an outcome yet provide little additional discrimination, particularly when the existing prediction model is already strong (Wald et al., 1999; Pepe et al., 2004; Cook, 2007). Conversely, a lack of predictive improvement does not establish that meaningful individual trajectories are absent: such trajectories may exist but be too poorly estimated to contribute useful information. Distinguishing these possibilities requires examining not only association and predictive performance, but also how reliably individual changes can be recovered from the available measurements.

Incident diabetes provides a natural setting in which to examine these issues. HbA1c (glycated haemoglobin) reflects average glycaemia over the preceding two to three months, the lifespan of erythrocytes being about 120 days (Cohen et al., 2008). It is measured repeatedly in several ageing cohorts, has an established diagnostic threshold of 48 mmol/mol (6.5%) (International Expert Committee, 2009; World Health Organization, 2011), and is strongly associated with diabetes risk as values approach that threshold (Zhang et al., 2010; Selvin et al., 2010; Tabák et al., 2012). Values of 42–47 mmol/mol (6.0–6.4%) are commonly described as prediabetes, or high risk of diabetes ^1^(International Expert Committee, 2009; National Institute for Health and Care Excellence, 2012). People in this range progress to diabetes considerably more often than those below it, although many do not progress for years or revert to normoglycaemia (Tabák et al., 2012). We used the English Longitudinal Study of Ageing (ELSA), a cohort in England, where HbA1c is measured according to a study schedule rather than in response to clinical suspicion, reducing bias from clinically driven testing. Previous studies have described average glycaemic trajectories retrospectively, aligned to the time of diabetes diagnosis (Tabák et al., 2009; Heianza et al., 2012), including an analysis of HbA1c in ELSA (Yu et al., 2023).

We address three questions. First, does longitudinal HbA1c history improve the prediction of incident diabetes beyond baseline HbA1c and conventional clinical risk factors (Lindström and Tuomilehto, 2003; Wilson et al., 2007; Hippisley-Cox and Coupland, 2017)? Second, is the rate of HbA1c change associated with incident diabetes after adjustment for baseline HbA1c, including when that level is estimated using previous measurements? Third, how reliably can individual HbA1c trajectories be estimated from sparsely sampled measurements in the presence of within-person variation?

To answer them, we compared a conventional clinical prediction model with models incorporating individual longitudinal features and with a self-supervised, time-aware Transformer trained on longitudinal histories (Tipirneni and Reddy, 2022). The features corresponded to the uses of history outlined above: deviation from the person’s usual level (the deviation claim), personal slope (the trajectory claim), change since the previous measurement, and the forecast probability of reaching the diagnostic threshold. We estimated their associations with incident diabetes and, to separate trajectory information from denoising, examined whether the slope association persisted after adjustment for a history-informed estimate of the current underlying HbA1c level. Finally, we modelled within-person variation as a serially correlated process (drift model) and used it to assess how reliably individual slopes can be estimated under different measurement schedules, varying the length of history and the frequency of measurement. The prediction models are not proposed for clinical use. The prediction analyses form a model development study with internal, participant-level cross-fitted validation; they ask whether personal HbA1c history adds information that could be used in repeated monitoring of older adults, not how well a specific model would perform in a new population.

## 2 Methods

### 2.1 Data

ELSA is a longitudinal study of adults aged 50 years and older living in England, with interviews conducted approximately every two years since 2002 and nurse-led health visits, including venous blood sampling, in selected waves (Steptoe et al., 2013). The sample is designed to represent adults aged 50 years and older living in private households in England. Analyses were unweighted, as the primary objectives were to estimate associations and assess predictive performance within the analytical cohort rather than to produce population-representative estimates. We used health-visit rounds 2 (2004–05), 4 (2008–09), 6 (2012–13), 8/9 (2016–2019; a single health-visit round spanning two waves) and 11 (2023–24) from the UK Data Service (SN 5050, End User Licence; Banks et al., 2026). Interview data were obtained from the Harmonized ELSA dataset (waves 1–10), produced by the Gateway to Global Aging Data and distributed with the ELSA data collection (Gateway to Global Aging Data, 2023), and from the wave-11 core interview file. HbA1c was recorded as a percentage in rounds 2 and 4, on the scale of the Diabetes Control and Complications Trial (DCCT), and in units of the International Federation of Clinical Chemistry and Laboratory Medicine (IFCC; mmol/mol) thereafter (Lloyd and Light, 2025). Measurement units were identified from the study documentation, and percentage values were converted to IFCC units using the IFCC–DCCT master equation, mmol/mol = (% *−* 2.15) *×* 10.929 (Hoelzel et al., 2004). Missing-value codes were removed, and a plausibility range of 15–200 mmol/mol was applied, which excluded fewer than 10 implausibly low values. Fasting glucose was measured only in participants who had fasted (as recorded at each health visit); because participants treated for diabetes were not asked to fast, its availability depended on diabetes status, and it was used to define diabetes but not as a predictor.

Systematic between-round differences in HbA1c, which would otherwise appear as within-person increases and inflate estimated slopes, were accounted for by round fixed effects in the mixed model used to estimate personal levels and slopes (Section 2.3); the estimated round effects, conditional on age and sex, were approximately +2 to +3 mmol/mol relative to round 2. These differences were already apparent between rounds 2 and 4, both recorded on the DCCT scale, and therefore cannot be explained by the subsequent change in measurement units. They may reflect changes in assay calibration, genuine calendar-time trends, or a combination of both.

### 2.2 Baseline visits and outcome

Diabetes was defined as the presence of any of the following: HbA1c *≥* 48 mmol/mol (*≥* 6.5%), fasting glucose *≥* 7.0 mmol/l (World Health Organization, 2011; International Expert Committee, 2009), a self-reported physician diagnosis, or use of diabetes medication. Laboratory measurements were obtained at health visits, whereas diagnoses and medication use were recorded at interviews; once present, diabetes was considered present at all later visits. A single measurement exceeding a diagnostic threshold was sufficient for classification in this study, although a clinical diagnosis would generally require confirmation. HbA1c was measured by an external laboratory in the blood sample taken at the health visit.

We introduce a *baseline visit* as a health visit at which the risk of developing diabetes by the next health visit was assessed. Baseline visits were drawn from rounds 4, 6 and 8/9. Eligibility required valid HbA1c, age and sex at the baseline visit; no diabetes by the above definition up to and including that visit; and at least one earlier HbA1c measurement obtained at least three months before the baseline visit while the participant was free of diabetes.

Incident diabetes was defined as meeting either laboratory criterion (HbA1c or fasting glucose) at the next health visit, or reporting a first physician diagnosis or diabetes medication use at any interview after the baseline visit, up to and including that of the next health-visit round. Diabetes types were not distinguished; type 1 diabetes can begin in adulthood but accounts for only a small share of diabetes diagnosed after the age of 30 (Thomas et al., 2018), so most incident cases were expected to be type 2. The main analysis included baseline visits with HbA1c available at the next health visit, so that the laboratory criterion could be assessed for all.

A participant could contribute more than one baseline visit, each representing a separate prediction using the information available at that time. Risk was therefore estimated per baseline visit rather than per participant. All visits from the same participant were assigned to the same cross-fitting fold to prevent information leakage (Section 2.3), and within-participant dependence was accounted for by participant-clustered standard errors and bootstrap intervals. The interval to the next health visit was approximately four years for rounds 4 and 6 and about seven years for round 8/9. Because the probability of observing incident diabetes increases with follow-up duration, all models included ln(years to next visit) as a covariate. The analysis therefore estimated the risk of diabetes detected by the next scheduled health visit, conditional on the interval between visits. Because the exact interval is not known at the baseline visit, this is an interval-adjusted prediction of diabetes detected at the next observed health visit rather than a fully prospective risk prediction. Detection or reporting of diabetes did not necessarily coincide with biological disease onset, and some cases may have been detected with delay.

### 2.3 Personal trajectories and longitudinal features

HbA1c trajectories were described by a linear mixed model (Laird and Ware, 1982; Verbeke and Molenberghs, 2000) fitted to non-diabetic visits:

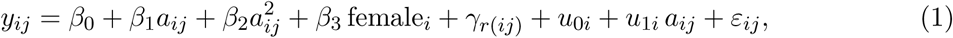

where *y_ij_* is the HbA1c value (mmol/mol) of person *i* at their *j*-th health visit, *a_ij_*= (age*_ij_ −*65)*/*10 is age centred at approximately the sample median and expressed in decades, female*_i_* indicates female sex (1 = female, 0 = male), and *γ_r_*_(_*_ij_*_)_ is the fixed effect of health-visit round *r*(*ij*), common to all participants measured in that round, with round 2 as reference (*γ*_2_ = 0). The fixed effects *β*_0_*,…,β*_3_ and *γ_r_* describe the population trajectory. The personal intercept *u*_0_*_i_* represents the person’s deviation from this trajectory at age 65, and the personal slope deviation *u*_1_*_i_* their deviation from the population age trend per decade; in mixed-model terms, both are random effects. We assumed (*u*_0_*_i_, u*_1_*_i_*)^T^ *∼ N* (0*, G*), a normal distribution with mean 0 and covariance matrix

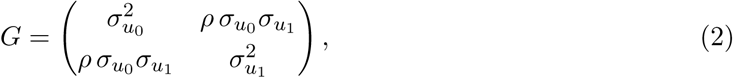

where *σ_u_*_0_ and *σ_u_*_1_ are the between-person standard deviations (SDs) of the intercept and slope and *ρ* their correlation, and *ε_ij_∼ N* (0*, σ*^2^) represents residual within-person variation between visits.

The analysis proceeded in two steps. First, the population parameters (the fixed effects, the covariance matrix *G* and the residual variance *σ*^2^) were estimated by restricted maximum likelihood (REML), the standard estimator for variance components (Patterson and Thompson, 1971; Harville, 1977). Second, for each baseline visit, the person’s intercept *u*_0_*_i_* and slope *u*_1_*_i_* were estimated by empirical Bayes: the population distribution *N* (0*, G*) served as a prior, which was updated with the person’s HbA1c measurements preceding the baseline visit, and the resulting posterior means were used as individual estimates (Robinson, 1991). To keep each participant’s data out of the parameters applied to them, we used cross-fitting (Chernozhukov et al., 2018): participants were split at random into five folds, a common choice that balances the amount of training data against computation, and the population parameters used for participants in one fold were estimated from the non-diabetic visits of the participants in the other four. Each participant was thus excluded from the estimation of the population parameters used to construct their features, while their individual estimates used only measurements preceding the baseline visit (for the forecast, also the HbA1c measured at the baseline visit itself).

Four features were derived per baseline visit (Figure 1). The first two correspond to the deviation and trajectory claims; the other two are simpler and combined summaries of the history:

- **Deviation from the expected value** (*z*): the baseline value minus the value predicted from the person’s earlier measurements, divided by the predictive SD of a new measurement.
- **Personal slope**: the posterior mean of the person’s rate of change at the reference age of 65 (population slope *β*_1_ plus the person-specific slope deviation *u*_1_*_i_*), in mmol/mol per decade. Because the population trajectory includes an age-squared term, this is the rate of change at age 65, not a constant lifetime slope. Differences in this feature between individuals therefore reflect their estimated deviations from the population slope. With a single earlier measurement, no within-person change is observed: the feature is then the population slope plus a small term proportional to the deviation of that measurement from the population trajectory, with a weight that depends on the age at which it was taken (Supplementary Section S3).
- **Change since last value**: the baseline value minus the last earlier value, after subtracting each round’s offset; the simplest summary of change, without a model.
- **Forecast**: the probability that the next HbA1c is *≥* 48 mmol/mol, from the posterior updated with the baseline value and projected over the interval to the next visit; it combines the current level and the personal trajectory.

**Figure 1:**
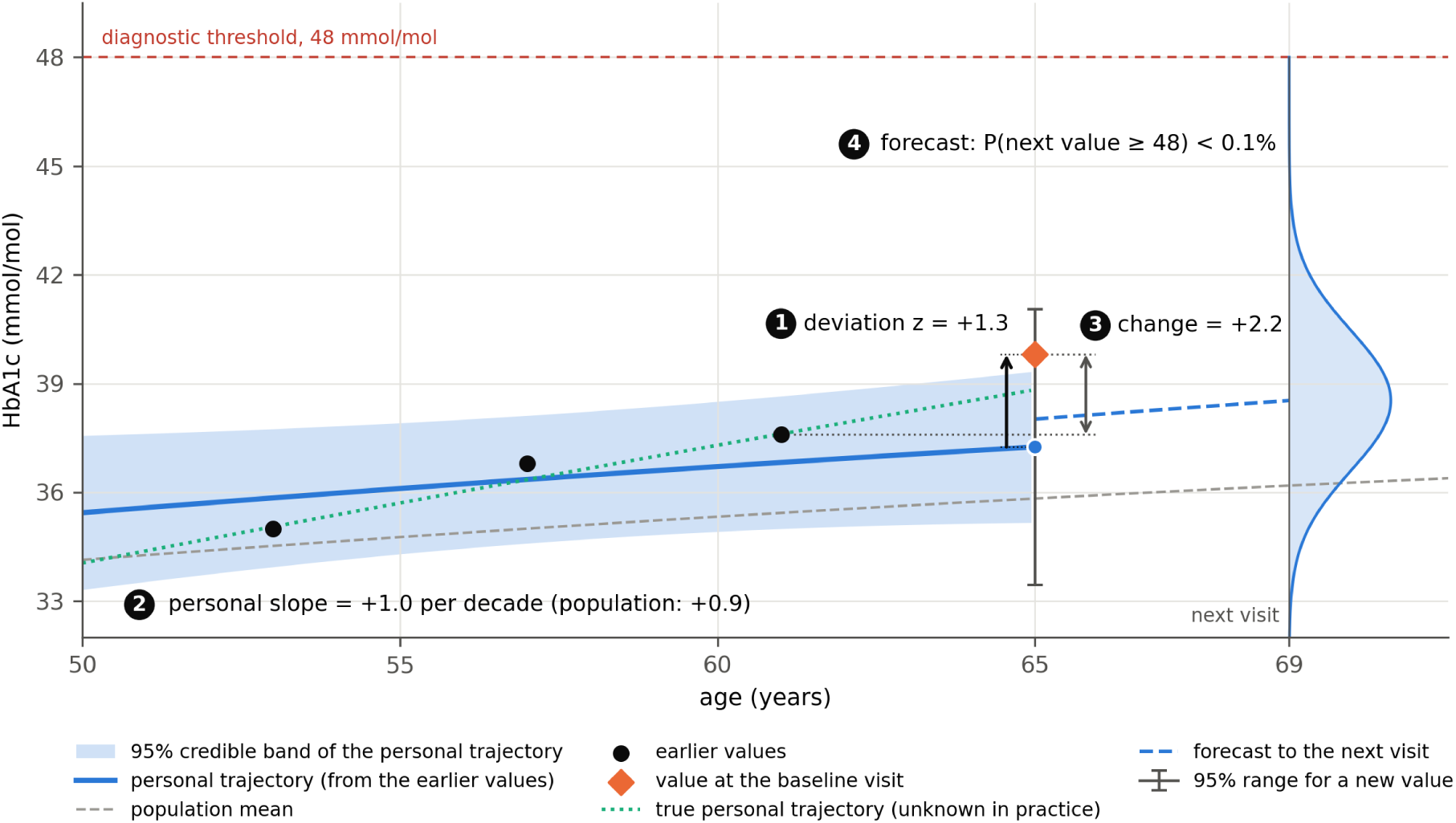
Construction of the four longitudinal features. Simulated HbA1c measurements for a hypothetical participant whose HbA1c rises faster than the population trajectory (true slope +3.0 vs +0.9 mmol/mol per decade), using model parameters estimated from ELSA (Table 2). The personal trajectory (blue line) and its 95% credible band are estimated from three earlier measurements (black dots); with so few values, the estimated slope (+1.0 per decade) is strongly shrunk towards the population slope and lies well below that of the true trajectory (green dotted line), which is unknown in practice. The grey dashed line represents the population mean, and the diamond marks the observed HbA1c at the baseline visit (age 65). (1) Deviation (*z*): the difference between observed and personally predicted HbA1c at the baseline visit (blue dot), scaled by the predictive SD for a new measurement (error bar: 95% prediction interval). (2) Personal slope: the rate of change of the estimated personal trajectory at age 65, compared with the population’s. (3) Change since the last measurement. (4) Forecast: the predicted distribution of HbA1c four years after the baseline visit (dashed line and density), after updating the personal trajectory with the baseline measurement, and the probability that it reaches the diagnostic threshold of 48 mmol/mol (red dashed line).

**Table 1:** Characteristics at the baseline visit, by incident diabetes at the next health visit.^a^.

|  | Analysis set (HbA1c at the next round) |  |  | Not in analysis: no HbA1c at next round<sup>b</sup> |
|---|---|---|---|---|
|  | All | Incident diabetes | No diabetes |  |
| Baseline visits, n | 5,828 | 234 | 5,594 | 4,387 |
| Persons, n | 3,603 | 234 | 3,466 | 4,278 |
| Age, years, mean (SD) | 67.7 (7.3) | 67.9 (6.9) | 67.7 (7.3) | 71.8 (9.2) |
| Women, n (%) | 3,321 (57.0) | 132 (56.4) | 3,189 (57.0) | 2,452 (55.9) |
| HbA1c, mmol/mol, mean (SD) | 38.7 (3.5) | 42.6 (3.5) | 38.6 (3.4) | 38.7 (3.6) |
| HbA1c, mmol/mol, median (IQR) | 39.0 (36.6–41.0) | 43.1 (40.0–45.0) | 38.8 (36.6–41.0) | 39.0 (36.6–41.0) |
| HbA1c, %, mean (SD)<sup>c</sup> | 5.70 (0.32) | 6.05 (0.32) | 5.68 (0.31) | 5.69 (0.33) |
| HbA1c 42–47.9 mmol/mol, n (%) | 1,194 (20.5) | 153 (65.4) | 1,041 (18.6) | 909 (20.7) |
| Personal slope, mmol/mol per decade, mean (SD)<sup>d</sup> | 0.94 (0.17) | 0.92 (0.21) | 0.95 (0.16) | 0.94 (0.19) |
| BMI, kg/m<sup>2</sup>, mean (SD) | 27.6 (4.7) | 30.7 (5.9) | 27.5 (4.5) | 27.4 (5.0) |
| Waist, cm, mean (SD)<sup>e</sup> | 95.0 (12.5) | 103.2 (13.2) | 94.7 (12.4) | 94.7 (13.0) |
| Systolic BP, mmHg, mean (SD) | 132.1 (17.0) | 136.5 (18.0) | 131.9 (16.9) | 133.3 (17.9) |
| BP medication, n (%) | 1,627 (28.2) | 103 (44.8) | 1,524 (27.5) | 1,530 (35.1) |
| HDL cholesterol, mmol/l, mean (SD) | 1.66 (0.45) | 1.50 (0.37) | 1.67 (0.45) | 1.64 (0.47) |
| Triglycerides, mmol/l, median (IQR) | 1.3 (0.9–1.8) | 1.7 (1.2–2.2) | 1.3 (0.9–1.8) | 1.3 (0.9–1.8) |
| Current smoker, n (%) | 506 (8.7) | 26 (11.2) | 480 (8.6) | 432 (9.9) |
| Vigorous activity weekly, n (%) | 2,213 (38.0) | 62 (26.5) | 2,151 (38.5) | 1,207 (27.5) |
| Fasting blood sample, n (%) | 4,302 (73.8) | 163 (69.7) | 4,139 (74.0) | 2,699 (61.5) |
| *Earlier HbA1c values, n (%)* |  |  |  |  |
| 1 | 3,552 (60.9) | 135 (57.7) | 3,417 (61.1) | 2,238 (51.0) |
| 2 | 1,815 (31.1) | 64 (27.4) | 1,751 (31.3) | 1,416 (32.3) |
| 3 | 461 (7.9) | 35 (15.0) | 426 (7.6) | 733 (16.7) |
| Years to next visit, median (IQR) | 4.2 (3.9–6.0) | 4.7 (4.0–6.9) | 4.2 (3.9–6.0) | 4.3 (4.0–6.2)<sup>f</sup> |
| *Missing, %* |  |  |  |  |
| BMI | 1.3 | 1.7 | 1.3 | 2.8 |
| Waist | 25.0 | 37.2 | 24.5 | 45.6 |
| Systolic BP | 4.3 | 6.4 | 4.2 | 6.6 |
| BP medication | 1.0 | 1.7 | 1.0 | 0.7 |
| HDL cholesterol | 0.6 | 1.3 | 0.6 | 0.6 |
| Triglycerides | 0.6 | 0.8 | 0.6 | 0.6 |
| Smoking | 0.3 | 0.4 | 0.3 | 0.3 |
| Physical activity | 0.0 | 0.0 | 0.0 | 0.0 |

<sup>a</sup> Except for persons, all figures refer to baseline visits; a person can contribute up to three baseline visits, and to more than one column. Percentages of categorical covariates refer to visits with non-missing values. BMI, body-mass index; BP, blood pressure; HDL, high-density lipoprotein; IQR, interquartile range; SD, standard deviation.
<sup>b</sup> Eligible baseline visits without HbA1c at the next round; not in the main analysis.
<sup>c</sup> HbA1c in % (DCCT/NGSP) = 0.09148× mmol/mol +2.152 (Hoelzel et al., 2004).
<sup>d</sup> Empirical Bayes estimates from the earlier values only, strongly shrunk towards the population slope (between-person SD 0.84 mmol/mol per decade; Section 3.1). Crude differences between columns partly reflect the dependence of the shrunk estimates on the person's level; the associations in Section 3.3 are adjusted for baseline HbA1c.
<sup>e</sup> Waist circumference was not available in the round 8/9 health-visit file.
<sup>f</sup> Among visits that had a next health visit, but without valid HbA1c.

**Table 2:**
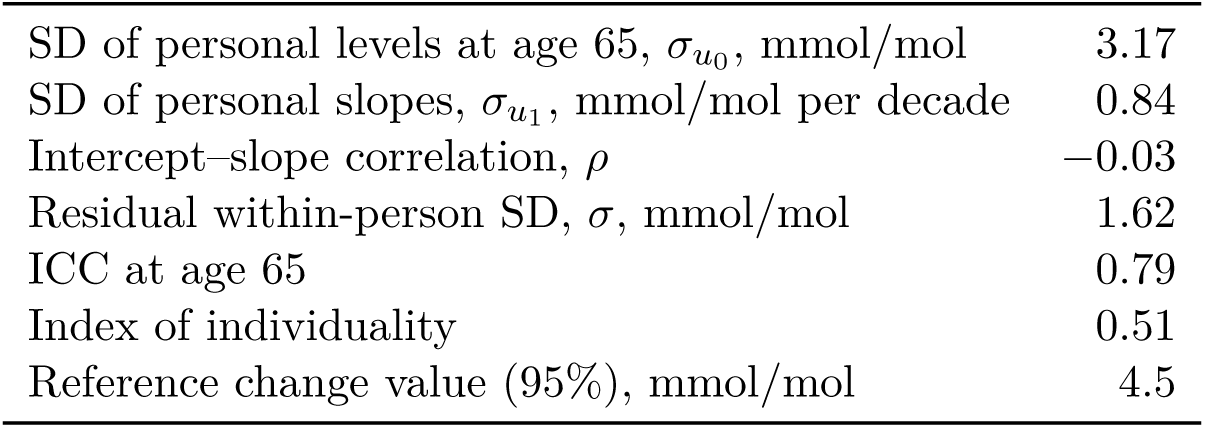
HbA1c variance components from the independent-noise trajectory model, equations (1) and (2) (mean over five folds), and the summaries of equation (3). The index of individuality and the reference change value assume independent within-person variation, an assumption relaxed by the drift model (Section 3.4).

|  |  |
|---|---|
| SD of personal levels at age 65, σ<sub>u<sub>0</sub></sub>, mmol/mol | 3.17 |
| SD of personal slopes, σ<sub>u<sub>1</sub></sub>, mmol/mol per decade | 0.84 |
| Intercept–slope correlation, ρ | −0.03 |
| Residual within-person SD, σ, mmol/mol | 1.62 |
| ICC at age 65 | 0.79 |
| Index of individuality | 0.51 |
| Reference change value (95%), mmol/mol | 4.5 |

**Table 3:** Out-of-fold discrimination of the prediction models (5,828 baseline visits, 234 cases). All models include ln(years to next visit); models C–E are model B plus the feature shown. Differences were computed on the same baseline visits in each bootstrap replicate, before rounding; 95% CIs from a participant-cluster bootstrap of the out-of-fold predictions with 1,000 replicates. Differences in subgroups are given in Supplementary Table S3.

| Model | AUC (95% CI) |
|---|---|
| A: HbA1c, age, sex | 0.819 (0.787–0.850) |
| B: A + clinical factors | 0.837 (0.806–0.868) |
| C: B + change since last value | 0.837 (0.807–0.868) |
| D: B + deviation *z* and personal slope | 0.835 (0.805–0.866) |
| E: B + logit of forecast probability | 0.837 (0.806–0.867) |
| T: Transformer (pretrained)<sup>a</sup> | 0.847 (0.817–0.876) |
| Difference | ΔAUC (95% CI) |
| B − A | +0.018 (0.005 to 0.031) |
| C − B | +0.001 (−0.001 to 0.002) |
| D − B | −0.002 (−0.005 to 0.002) |
| E − B | 0.000 (−0.005 to 0.004) |
| T − B | +0.010 (−0.006 to 0.026) |

<sup>a</sup> Also received variables not in model B, including waist circumference, further laboratory markers and self-rated health (Section 2.7); alternative configurations in Supplementary Table S6.

Finally, as conventional summaries of between-and within-person variation under model (1), assuming independent residual variation (Harris, 1974; Fraser and Harris, 1989; Fraser, 2001), we calculated three measures:

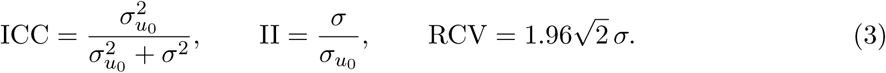

The intraclass correlation (ICC) is the share of variance attributable to stable differences between individuals, and the index of individuality (II) compares within-person with between-person variability; values of the II below approximately 0.6 suggest that population reference intervals may be of limited use for interpreting individual measurements. Both use the between-person variance at the reference age of 65, *σ*^2^; because people differ in their slopes, between-person variance changes with age. The reference change value (RCV) depends only on the within-person variance *σ*^2^ and gives the minimum difference between two measurements from the same individual that exceeds visit-to-visit variation at the 95% level. The drift model (Section 2.8) relaxes the assumption of independent residual variation on which the II and the RCV rely.

The distributional assumptions of model (1), fitted once to all non-diabetic visits for this purpose, were checked with normal quantile–quantile plots of the standardised conditional residuals and of the empirical Bayes estimates of personal levels and slopes, and with plots of residuals against fitted values (Supplementary Figure S1). The assumption of independent within-person residuals was examined by the correlation of residuals between pairs of health-visit rounds (Supplementary Figure S2).

### 2.4 Prediction analyses

We examined whether longitudinal HbA1c features improved the prediction of incident diabetes beyond baseline HbA1c and conventional clinical risk factors. We compared five logistic regression models, beginning with a minimal model (A), extending it with clinical risk factors (B), and then adding alternative representations of personal HbA1c history (C–E). All models included ln(years to next visit) to account for differences in follow-up duration:

- Model A: baseline HbA1c, age and sex;
- Model B: Model A plus body-mass index (BMI), systolic blood pressure, high-density lipoprotein (HDL) cholesterol, log triglycerides, smoking, weekly vigorous activity and blood-pressure medication;
- Model C: Model B plus change since the last HbA1c measurement;
- Model D: Model B plus deviation *z* and personal slope;
- Model E: Model B plus the logit of the forecast, i.e. the predicted probability that HbA1c reaches 48 mmol/mol at the next health visit (the forecast concerns HbA1c only, whereas the outcome also includes glucose, reported diagnoses and medication).

Model B included clinical predictors commonly used in established diabetes risk scores (Lindström and Tuomilehto, 2003; Wilson et al., 2007; Hippisley-Cox and Coupland, 2017; Noble et al., 2011; Abbasi et al., 2012) and available in ELSA; their sources and timing are described in Supplementary Section S2. The models were fitted on standardised predictors with a fixed L2 penalty to stabilise the estimates, maximising 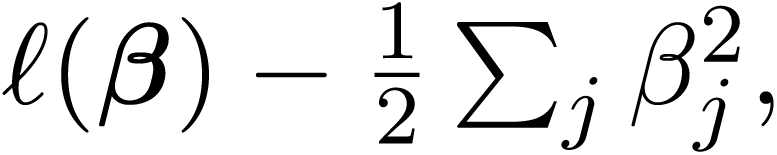 where *ℓ* is the log-likelihood summed over baseline visits and the sum runs over the coefficients of the predictors (the intercept was not penalised); the penalty strength was not tuned and was the same in all models. Missing clinical covariates were imputed with the median, with an additional missingness indicator for each covariate with missing values (Sperrin et al., 2020). Predictions were generated out of fold using the same participant-level folds as for the longitudinal features: imputation medians, missingness indicators and standardisation parameters were estimated within each training fold and applied unchanged to the held-out participants, and the prediction models were fitted only to the training participants of each fold. The sample size was determined by the available eligible ELSA observations rather than by a prospective sample-size calculation; the relatively small number of incident cases limits the precision of the performance estimates. Final versions of models A and B, refitted to all baseline visits, are specified in Supplementary Section S2 and Table S4; models C–E are specified in the analysis code.

Performance metrics weighted each baseline visit equally, rather than each participant. We assessed discrimination (area under the receiver operating characteristic curve, AUC), calibration (intercept and slope from a logistic regression of the outcome on the logit of the predicted risk), and accuracy of predicted probabilities (Brier score). To examine the practical implications for screening, we also compared how many baseline visits followed by incident diabetes were identified when each model selected the highest-risk 5% or 10% of baseline visits; because participants could contribute more than one baseline visit, these counts refer to visits rather than individuals. Confidence intervals (CIs; 95%) for AUCs, AUC differences and differences in screening counts were obtained from a participant-cluster bootstrap of the out-of-fold predictions with 1,000 replicates (Efron and Tibshirani, 1993; Field and Welsh, 2007); they are conditional on these predictions and do not include the uncertainty from refitting the models. Differences in discrimination were also estimated in subgroups of baseline visits, to examine whether personal history adds information where baseline HbA1c is least informative or where more history is available: visits with HbA1c *<* 42 mmol/mol, visits with one and with at least two earlier HbA1c values, and visits with an approximately four-year interval to the next visit (rounds 4 and 6).

### 2.5 Association analyses

We estimated the association of each longitudinal feature with incident diabetes, adjusted for baseline HbA1c and for the established clinical risk factors of Model B, which may be related to both HbA1c change and diabetes risk.

For baseline visit *k* with feature *e_k_*, standardised to mean 0 and SD 1, we fitted

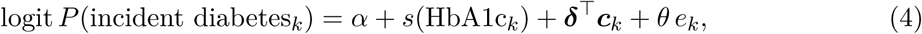

where *s*(*·*) is a natural cubic spline in baseline HbA1c (4 degrees of freedom; boundary knots at the minimum and maximum, interior knots at the quartiles; Durrleman and Simon, 1989; Harrell, 2015), ***c****_k_* contains the other covariates of Model B (age, sex and clinical risk factors) and ln(years to next visit), with coefficient vector ***δ***, and *e^θ^* is the odds ratio (OR) per SD of the feature at the same HbA1c and covariate values. A flexible term for HbA1c was used because risk rises nonlinearly as HbA1c approaches the diagnostic threshold (Zhang et al., 2010; Selvin et al., 2010), and the longitudinal features, being derived from the same HbA1c series, may be correlated with baseline HbA1c; with a linear term, part of this nonlinearity could be attributed to the features. These models were fitted without penalty, with missing covariates imputed with the median and a missingness indicator, and standard errors clustered by participant (Liang and Zeger, 1986). For comparison, we also report the slope association without adjustment and adjusted only for follow-up duration, ln(years to next visit).

We repeated these analyses separately for baseline visits below and within the prediabetes range (HbA1c *<* 42 and 42–47.9 mmol/mol; International Expert Committee, 2009), where identifying differences in risk may be particularly relevant. Because a personal slope reflects observed within-person change only when at least two earlier HbA1c measurements are available, we report the slope association among these baseline visits alongside that in all baseline visits. With a single earlier value, the estimated slope is determined by the deviation of that value from the population trajectory rather than by observed change (Section 2.3; Supplementary Section S3). We also tested whether the slope association varied with baseline HbA1c, using an interaction between the personal slope and HbA1c range (*<* 42 versus 42–47.9 mmol/mol) in a single model including all baseline visits, with covariate effects common to both ranges; the ratio of the slope ORs in the two ranges, *e^θ×^* for interaction coefficient *θ*_×_, provides a test and CI that separate subgroup models do not, because the same participant can contribute baseline visits to both ranges. Each feature was standardised by its SD within the analysed set of baseline visits, so that ORs in subgroups refer to the subgroup-specific SD; the interaction model used the SD in all baseline visits. As a sensitivity analysis, subgroup associations were also expressed per SD in all baseline visits, so that they refer to the same absolute increment in the feature.

An association between the personal slope and incident diabetes after adjustment for observed baseline HbA1c could arise without the rate of change carrying independent information. A single measurement represents the underlying current HbA1c level imperfectly, and when within-person deviations persist over several years (slow drift; Section 2.8), earlier measurements may also provide information about that level. An estimated slope could therefore be associated with diabetes partly because it provides information about the current level not captured by the baseline measurement. To examine this possibility, we refitted model (4) for the personal slope, replacing observed baseline HbA1c with a history-informed estimate of the underlying current level obtained from the drift model (Section 2.8).

### 2.6 Sensitivity analyses

Unless stated otherwise, sensitivity analyses concerned the association of the personal slope with incident diabetes (Supplementary Section S3).

We examined sensitivity to slope estimation and adjustment for recent HbA1c level. In the primary analysis, personal slopes were estimated using only measurements preceding the baseline visit and therefore described the trajectory before the HbA1c measurement used for adjustment. To assess the effect of incorporating the most recent observation, we repeated the analysis using slopes estimated from measurements up to and including baseline, while retaining adjustment for baseline HbA1c. In this analysis, the slope and the adjustment variable shared the baseline measurement. As an alternative adjustment for recent HbA1c level, we used the mean of the baseline and immediately preceding measurements, correcting the earlier value for the difference between round effects.

We also examined sensitivity to outcome definition and follow-up duration. We defined incident diabetes using only HbA1c *≥* 48 mmol/mol at the next health visit. For this analysis, we excluded baseline visits for which diabetes at follow-up was identified only through fasting glucose, self-reported diagnosis or medication use, because treatment may have lowered follow-up HbA1c below the diagnostic threshold. Because this exclusion depended on follow-up information, we also repeated the analysis with these visits classified as non-cases. Separately, we restricted the sample to baseline visits in rounds 4 and 6, which had an approximately four-year interval to the next health visit, to reduce variation in follow-up duration.

To assess sensitivity to missing covariates, we repeated the association analyses for all four longitudinal features (Section 2.3) using multiple imputation instead of median imputation (Rubin, 1987; White et al., 2011; van Buuren, 2018). To address selective follow-up, we weighted the analysis by the inverse probability of having an HbA1c measurement at the next health visit (Seaman and White, 2013). In a separate analysis, we extended outcome ascertainment to include baseline visits for which diabetes status at follow-up was known from interviews. For participants who died before the next health visit, we conducted a scenario-based analysis that included their baseline visits with hypothetical diabetes outcomes generated from prediction models using baseline HbA1c and clinical covariates, either including or excluding the personal slope.

Finally, we also examined the potential influence of participation history. Participants with at least two earlier HbA1c measurements had generally remained in the study longer; within this subgroup, we therefore modelled age with a natural cubic spline instead of a linear term, or additionally adjusted for birth cohort or for length of participation, one at a time.

### 2.7 Time-aware Transformer

To examine whether a flexible model could extract predictive information from longitudinal histories beyond that captured by the engineered features, we used a self-supervised time-aware Transformer for sparse, irregularly sampled data (STraTS; Tipirneni and Reddy, 2022). The model represented each participant’s observations up to and including the baseline visit as (time, variable, value) triples covering 31 health-visit and interview variables. Within each training fold, the network was pretrained on all ELSA participants with at least two observation times, not only the analysis cohort, to forecast the first value of each variable within 2.5 years after a randomly chosen observation time, a window fixed in advance to cover the next biennial interview wave. It was then fine-tuned to predict incident diabetes. Test-fold participants were excluded from pretraining, fine-tuning and normalisation. An otherwise identical network trained without pretraining served as an ablation.

Both networks were evaluated using the same participant-level folds as the logistic regression models and compared with Model B using paired participant-cluster bootstrap intervals for differences in AUC. Because the Transformer received additional variables, including waist circumference, further laboratory markers and self-rated health, the comparison assessed whether a flexible model with access to the broader longitudinal record improved prediction, rather than isolating the effect of the architecture. Because fine-tuning weighted cases by the ratio of non-cases to cases, which shifts predicted risks upwards, the Transformer was compared with the logistic models by discrimination only; its predicted risks were not recalibrated. Three alternative configurations, varying model dimension or the number of encoder layers, served as robustness checks. Architecture, pretraining and optimisation details are provided in Supplementary Section S2 (Table S5).

### 2.8 Drift model

A persistent increase in a person’s HbA1c may be difficult to distinguish from a fluctuation that lasts several years. The mixed model (1) assumes that deviations from the personal trajectory are independent from visit to visit. If deviations persist, measurements close in time are more similar than distant ones (serial correlation), and a temporary rise can resemble a personal slope. To allow for this, we added a stationary Ornstein–Uhlenbeck (OU) process to the within-person variation (Uhlenbeck and Ornstein, 1930; Diggle, 1988; Diggle et al., 2002). For person *i* with HbA1c values ***y****_i_* measured at ages *t_i_*_1_*,…,t_ini_*,

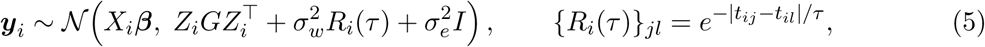

where the rows of *X_i_* contain the fixed-effect terms of model (1) at each visit, so that *X_i_**β***(with ***β*** including the round effects *γ_r_*) is the population trajectory; the rows of *Z_i_* are (1*, a_ij_*), so that *Z_i_GZ_i_*^T^ is the covariance of the personal intercepts and slopes of model (1); and *I* is the identity matrix. The term σ*_w_*^2^*R_i_* (*t*), which we call *drift*, describes deviations that persist for years but return towards the personal trajectory: drift values Δ years apart have correlation *e*^−Δ^*^/τ^*, so the time constant *τ* sets how long they last. The term σ_e_^2^ *I* represents independent variation, including measurement error. The fraction of within-person variance due to drift is π=σ*_w_*^2^/(σ*_w_*^2^+σ*_e_*^2^); with σ*_w_*^2^=0, the model reduces to model (1) with independent noise (Figure 2).

**Figure 2:**
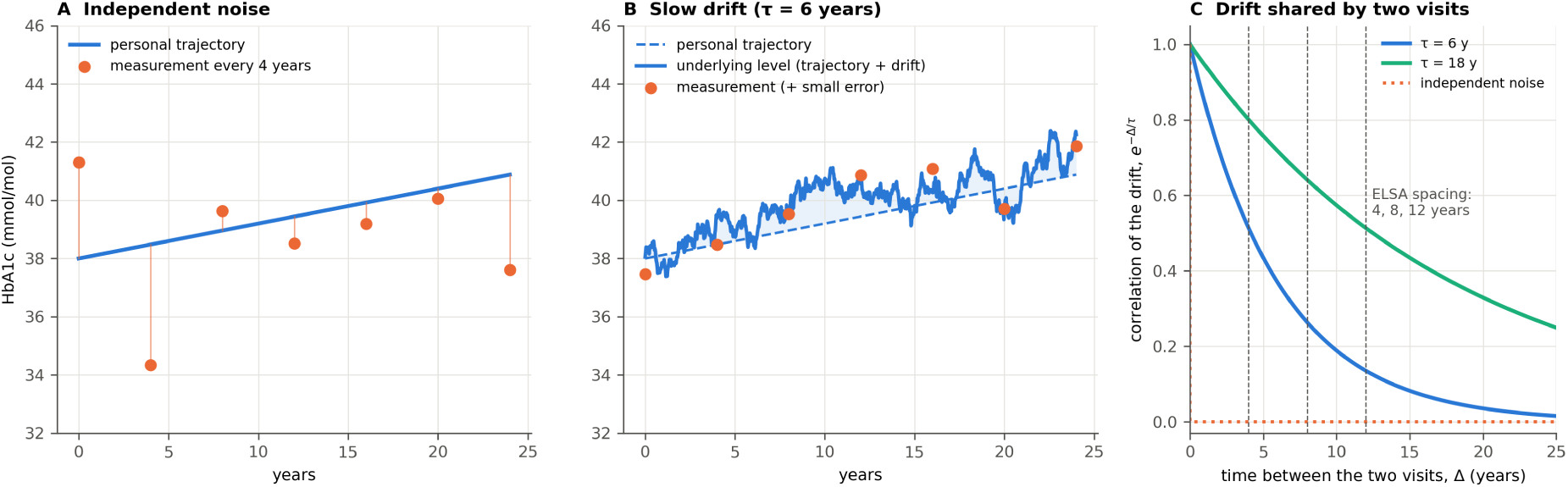
Independent noise and drift (simulated values). (A) Independent noise: measurements scatter around the personal trajectory, each independently of the others. (B) Ornstein–Uhlenbeck drift with *τ* = 6 years: the underlying level (solid) departs from the personal trajectory (dashed) in spells lasting years, and measurements follow it. (C) Correlation of the drift between two visits as a function of the time between them, *e^−^*^Δ^*^/τ^* for two different time constants; dashed lines mark ELSA’s spacing of 4, 8 and 12 years; the dotted line shows independent noise, which carries no correlation between visits.

Slopes and drift differ in how differences between a person’s measurements grow with the time between them. For two measurements Δ *>* 0 years apart, half the expected squared difference of their deviations from the population trajectory (the semivariogram; Diggle, 1988; Diggle et al., 2002) is

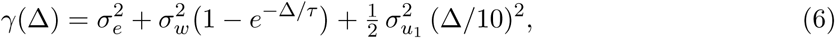

with Δ*/*10 in decades, the unit of the slope. The drift term levels off once Δ is several times *τ*, whereas the slope term keeps growing. Persistent slopes can therefore be separated from drift only through intervals that are long relative to *τ*; additional measurements at short intervals mainly inform the noise and drift components.

Restricting the model to non-diabetic visits may truncate rising trajectories: after HbA1c crosses the diagnostic threshold, subsequent measurements are excluded, so a persistent increase can appear temporary (Hernán et al., 2004; Tobin et al., 2005). We therefore fitted two additional versions of the model, each including the same non-diabetic visits plus each participant’s first HbA1c value *≥* 48 mmol/mol (the threshold-crossing value). The first used the observed crossing values. Because these values may reflect an accelerating rise before diagnosis (Tabák et al., 2009; Heianza et al., 2012), which a stationary drift process does not describe, the second used only the information that the threshold had been crossed, treating each crossing value as censored at 48 mmol/mol: instead of the density of its measured value, the participant’s likelihood includes the model probability that HbA1c was at least 48 mmol/mol at that visit, given their earlier values (Supplementary Section S4).

For each fit, we estimated six covariance parameters: the intercept and slope SDs and their correlation, *σ_w_*, *σ_e_* and *τ*. Estimation used restricted maximum likelihood (REML), except for the censored fit, which used maximum likelihood. Because the data contain little information on *τ*, we examined its profile likelihood and obtained an approximate 95% interval (Venzon and Moolgavkar, 1988; Pawitan, 2001). We compared the drift model with the independent-noise model using the likelihood-ratio statistic and the Akaike information criterion (AIC; Akaike, 1974). Further details are given in Supplementary Section S4.

To separate trajectory information from denoising, the drift model also provided an estimate of each person’s underlying current HbA1c level for the association analysis (Section 2.5 and Figure 2). We defined this level as the personal trajectory plus drift at the baseline visit, excluding the independent component, and estimated it by its conditional mean given the person’s measurements up to and including that visit. Because the personal trajectory and drift persist over years, earlier measurements contribute to this estimate. For the two fits including crossing values, the model parameters were estimated once using all participants; association analyses using these estimates therefore did not use an out-of-sample adjustment.

Finally, we assessed whether adjustment for the estimated underlying current level could reduce an apparent slope association caused by imperfect measurement of the current level. We simulated HbA1c histories for 12,000 hypothetical participants from a drift model with parameters close to those fitted with the crossing values, with values about four years apart, excluding participants with any value *≥* 48 mmol/mol. Incident diabetes was generated from the underlying current level alone, so that the personal slope carried no information of its own, and the slope association was then estimated with adjustment for either the observed last value or the estimated level (Supplementary Section S4).

### 2.9 Observability of personal slopes

Over short periods, persistent drift can look like a personal slope (Equation (6)), so how well the two can be separated depends on the number, spacing and duration of measurements. The fitted trajectory models (Equation (5)) allow us to quantify how reliably personal slopes can be recovered under different measurement schedules. We applied the calculation to the independent-noise model, the special case σ*_w_*^2^=0, and to the two drift fits including threshold-crossing values (as observed and censored at 48 mmol/mol). We considered hypothetical schedules with measurements every 0.25 to 4 years over histories of 4 to 30 years. The shortest interval, three months, corresponds to the usual minimum interval for repeat HbA1c testing: because HbA1c reflects glycaemia over the lifespan of erythrocytes, more frequent measurements would largely share the same glycaemic history (Cohen et al., 2008).

For each schedule, we asked how much the measurements reduce the uncertainty about a person’s slope, an approach used to compare longitudinal designs (Willett, 1989; Raudenbush and Liu, 2001; Rast and Hofer, 2014; Brandmaier et al., 2018). Before any measurement, this uncertainty is the between-person slope variance, *G*_22_. After the measurements, it is the posterior variance Var(*u*_1_ *|* ***y***). To compute it, consider a hypothetical person with intercept and slope ***u*** = (*u*_0_*, u*_1_), measured at regular intervals over the observation period with the last measurement at age 65, the reference age of the personal intercept; ***y*** denotes these measurements, and the subscript *i* is omitted because no participant’s data are involved. The posterior covariance matrix of ***u*** is (Robinson, 1991; Verbeke and Molenberghs, 2000)

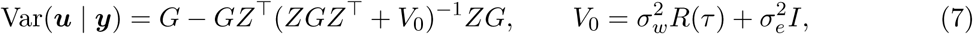

where *Z* is the matrix with rows (1*, a_j_*) for the measurement ages and *R*(*τ*) is defined as in (5); Var(*u*_1_ *|* ***y***) is its second diagonal element. In equation (7), *ZGZ*^T^ represents variation arising from personal intercepts and slopes, while *V*_0_ represents drift and independent noise. Conditional on the fitted covariance parameters, the posterior variance depends only on the measurement schedule, not on the observed HbA1c values, and can therefore be calculated without simulating measurements. Independent fluctuations can be averaged out through repeated measurements, whereas serially correlated drift reduces the additional information provided by closely spaced observations (Diggle et al., 2002). We defined slope reliability as 1 *−* Var(*u*_1_ *|* ***y***)*/G*_22_, the fraction of between-person slope variance resolved by the measurements, ranging from 0 (no information) to 1 (perfect recovery). Under the Gaussian model, this equals the squared correlation between the estimated slope *û*_1_ = *E*(*u*_1_ *|* ***y***) and the underlying slope *u*_1_ (Willett, 1989; Raudenbush and Liu, 2001), so we compute

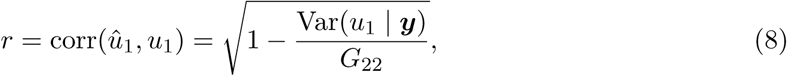

Because *û*_1_ is computed from the full covariance matrix *G*, *r* also depends on the intercept– slope correlation *ρ*. At *ρ* = 1, the slope is a fixed multiple of the intercept, *u*_1_ = *c u*_0_ with 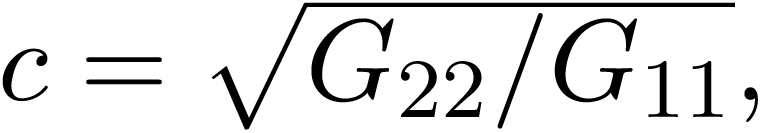 so that *û*_1_ = c *û*_0_ and *r* = corr(*û*_0_, *û*_0_): the correlation for the slopes then equals how well the person’s level is estimated. Even a single measurement would then give a high *r*, although it contains no information on change over time. We therefore calculated *r* with *ρ* set to zero and the other fitted covariance parameters held fixed, so that *û*_1_ no longer draws on information transferred through the assumed correlation between the personal level at age 65 and the slope. This is a conditional calculation, not a separately fitted model. Its purpose is to examine the recoverability of personal change without information transferred from the personal intercept through the assumed correlation. Results with the fitted correlation are given in Supplementary Table S12.

### 2.10 Reporting

Reporting follows STROBE for the association analyses (von Elm et al., 2007) and TRIPOD+AI for the prediction analyses (Collins et al., 2024); the completed checklists are given in Supplementary Tables S13 and S14.

## 3 Results

### 3.1 Cohort, measurement histories and HbA1c variation

Of 10,215 eligible baseline visits identified among 30,958 health-visit records in rounds 2 to 8/9, 5,828 visits from 3,603 participants had HbA1c measured at the next health visit and formed the analysis set (Figure 3). In total, 234 baseline visits (4.0%) were followed by incident diabetes over a median follow-up of 4.2 years. Mean age at baseline was 67.7 years, 57.0% of visits were from women, 98.5% of participants were of White ethnicity, and 20.5% had HbA1c of 42–47.9 mmol/mol (Table 1). Visits followed by incident diabetes had higher baseline HbA1c, BMI and waist circumference, more frequent use of blood-pressure medication, and less frequent weekly vigorous activity. Characteristics by HbA1c range are given in Supplementary Table S1.

**Figure 3:**
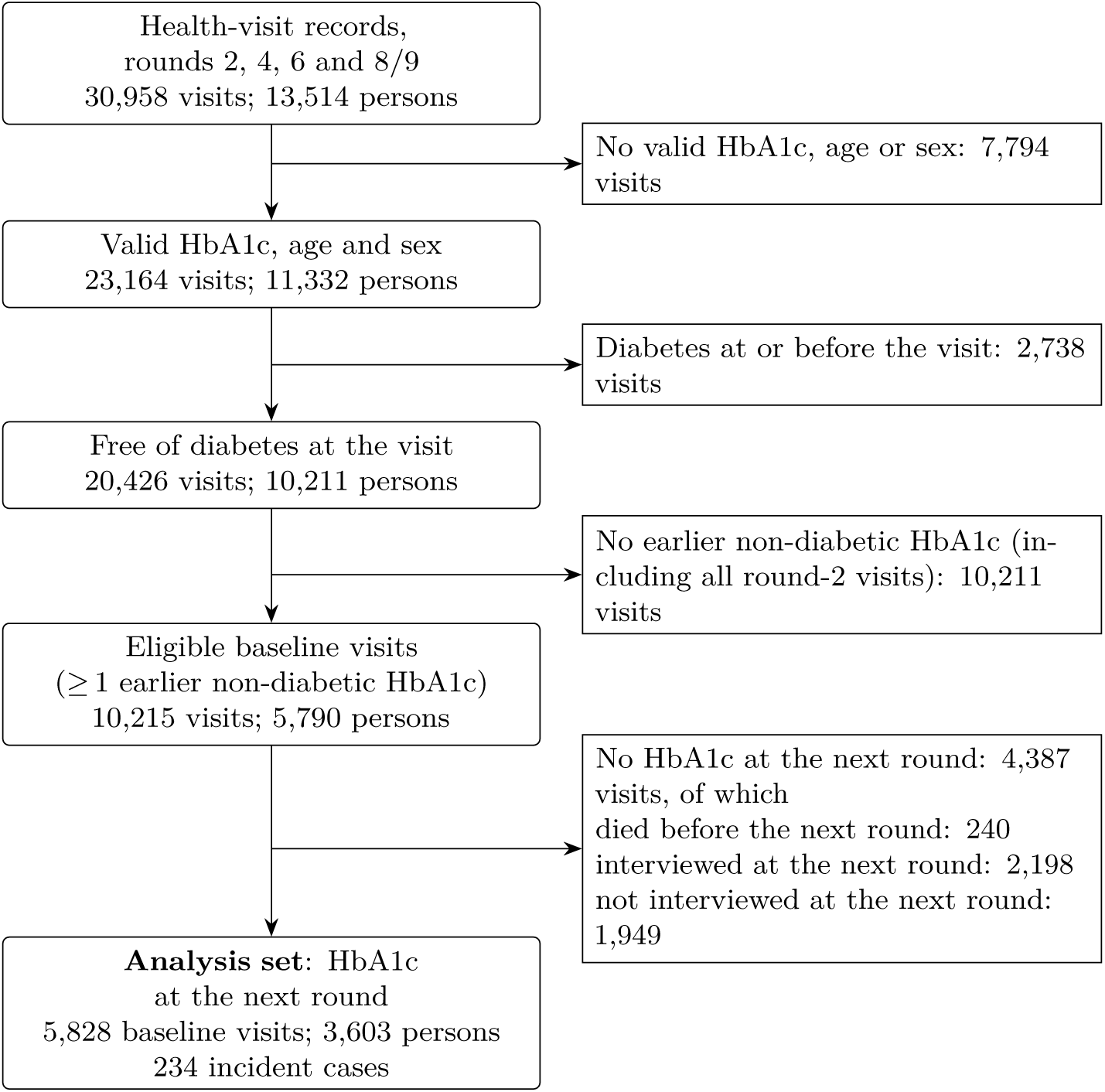
Participant flow. A participant can contribute up to three baseline visits (rounds 4, 6 and 8/9). Round-11 health visits cannot be baseline visits, as no later round exists; they provide the outcome for round-8/9 baseline visits and enter the trajectory and drift models (Table 4). Baseline visits without HbA1c at the next round were used for the weights for loss to follow-up. Those for which the diabetes question was answered at an interview in the next round, or diabetes was reported at an interview between the baseline visit and the next round, formed the extended outcome set (Supplementary Table S9); because of these reports, the set also includes some visits shown here as died or not interviewed at the next round. Visits ending in death were also added back in a separate sensitivity analysis (Section 2.6).

**Table 4:** Independent-noise and drift models of within-person HbA1c variation, fitted to three data sets: non-diabetic visits only, plus each participant’s threshold-crossing value, and with that value censored at 48 mmol/mol.

|  | Non-diabetic visits | Plus crossing value<br>(HbA1c ≥ 48) | Crossing value<br>censored at 48 |
|---|---|---|---|
| Participants; visits | 11,396; 23,599 | 11,396; 24,000 | 11,396; 24,000 |
| Slope SD, independent noise | 0.84 | 1.11 | 1.03 |
| Slope SD, drift model | 0.06 | 0.81 | 0.33 |
| Drift share *π* | 0.79 | 0.81 | 0.77 |
| Drift time constant *τ*, years | 50 (12.5–87.6) | 5.8 (4.3–7.9) | 26 (14.4–43.2) |
| Likelihood ratio vs independent noise | 144 | 382 | 232 |
| ΔAIC | −140 | −378 | −228 |

Slope SD, between-person SD of personal slopes (mmol/mol per decade); *π*, share of residual within-person variance due to drift; *τ* with 95% profile-likelihood interval; ΔAIC = AIC(drift) − AIC(independent noise), negative values favouring drift. Fitted by REML, the censored fit by maximum likelihood (ML). Visits from all health-visit rounds (2–11), including round 11 (Figure 3). Fits with capped crossing values in Supplementary Table S11.

HbA1c was unavailable at the next health visit for 4,387 eligible baseline visits. Compared with visits in the analysis set, these visits were from older participants (mean age 71.8 versus 67.7 years), with more frequent use of blood-pressure medication (35% versus 28%), but similar mean baseline HbA1c and estimated personal slopes (Table 1).

The histories available for estimating personal trajectories were sparse: 60.9% of baseline visits had only one earlier HbA1c measurement, 31.1% had two, and 7.9% had three. Consecutive measurements were a median of 4.0 years apart, and the first earlier measurement was a median of 4.3 years (IQR 3.9–7.9) before baseline (Supplementary Table S2). Because the primary slopes used only measurements before baseline, observed within-person change informed them for only 39.1% of visits.

Under the independent-noise trajectory model (Equation (1)), between-person variation in HbA1c at age 65 was greater than residual within-person variation (SDs 3.17 and 1.62 mmol/mol, respectively), giving an ICC at age 65 of 0.79 and an index of individuality of 0.51 (Table 2). The corresponding reference change value was 4.5 mmol/mol: a difference between two measurements had to exceed this to be distinguishable from fluctuation around the personal trajectory. The estimated between-person SD of personal slopes was 0.84 mmol/mol per decade, whereas the empirical Bayes slope estimates derived from the available histories had an SD of only 0.17 mmol/mol per decade, reflecting strong shrinkage towards the population slope.

Model diagnostics indicated departures from the assumed residual distribution and temporal structure. Standardised conditional residuals were approximately symmetric (skewness 0.02) but more peaked and heavier-tailed than a normal distribution (excess kurtosis 2.4). Empirical Bayes estimates of personal levels were approximately normal apart from the lower tail (Supplementary Figure S1). Residual correlations between rounds declined with increasing time between measurements more rapidly than implied by the fitted independent-noise model (Supplementary Figure S2), motivating the drift analysis reported in Section 3.4.

### 3.2 Limited incremental predictive value of personal HbA1c history

Personal HbA1c history added little discrimination beyond baseline HbA1c and clinical risk factors (Table 3). Adding clinical factors to baseline HbA1c, age and sex improved the AUC from 0.819 to 0.837 (B *−* A: ΔAUC = +0.018, 95% CI 0.005 to 0.031). The improvement was numerically larger among visits with HbA1c *<* 42 mmol/mol (+0.049, 95% CI 0.020 to 0.080;

Supplementary Table S3). Adding personal-history features to the clinical model produced little further change: differences in AUC were small in the full analysis set and in every subgroup examined, including visits with at least two earlier HbA1c measurements (C *−* B, D *−* B and E *−* B: all *|*ΔAUC*| ≤* 0.006, with all 95% CIs including zero).

Personal-history features also produced little change in screening performance. When the highest-risk 5% of baseline visits were flagged, model B identified 83 of the 234 visits followed by incident diabetes (35.5%), compared with 84 for model D, which added deviation and personal slope (difference +1, 95% CI *−*5 to 9), and 85 for model E, which added the forecast (+2, 95% CI *−*4 to 10). At a screening fraction of 10%, all three models identified 135 visits followed by incident diabetes (57.7%). Across logistic models A–E, calibration slopes ranged from 0.93 to 1.00 and calibration intercepts from *−*0.17 to 0.00; Brier scores ranged from 0.033 to 0.034.

The pretrained Transformer achieved an AUC of 0.847 (95% CI 0.817 to 0.876), compared with 0.837 for model B (ΔAUC = +0.010, 95% CI *−*0.006 to 0.026). Across three alternative Transformer configurations, differences from model B ranged from *−*0.009 to *−*0.001, with all 95% CIs including zero (Supplementary Table S6). In the pretraining task, forecast error averaged across variables was 16–18% lower than with last-observation-carried-forward prediction. For HbA1c, however, forecast error ranged from 5% lower to 9% higher across folds. Pretraining also produced little change in diabetes discrimination compared with random initialisation (pretrained minus randomly initialised: ΔAUC = *−*0.002, 95% CI *−*0.014 to 0.009).

### 3.3 Association of personal HbA1c slopes with incident diabetes

Personal HbA1c slopes were associated with incident diabetes after adjustment for baseline HbA1c and clinical factors, despite adding little discrimination (OR per SD 1.18, 95% CI 1.02– 1.36; Figure 4A, Supplementary Table S7). Among baseline visits with at least two earlier measurements, for which slope estimation incorporated observed within-person change, the OR was 1.39 (1.11–1.74). The other personal-history features showed no clear association (Figure 4A).

**Figure 4:**
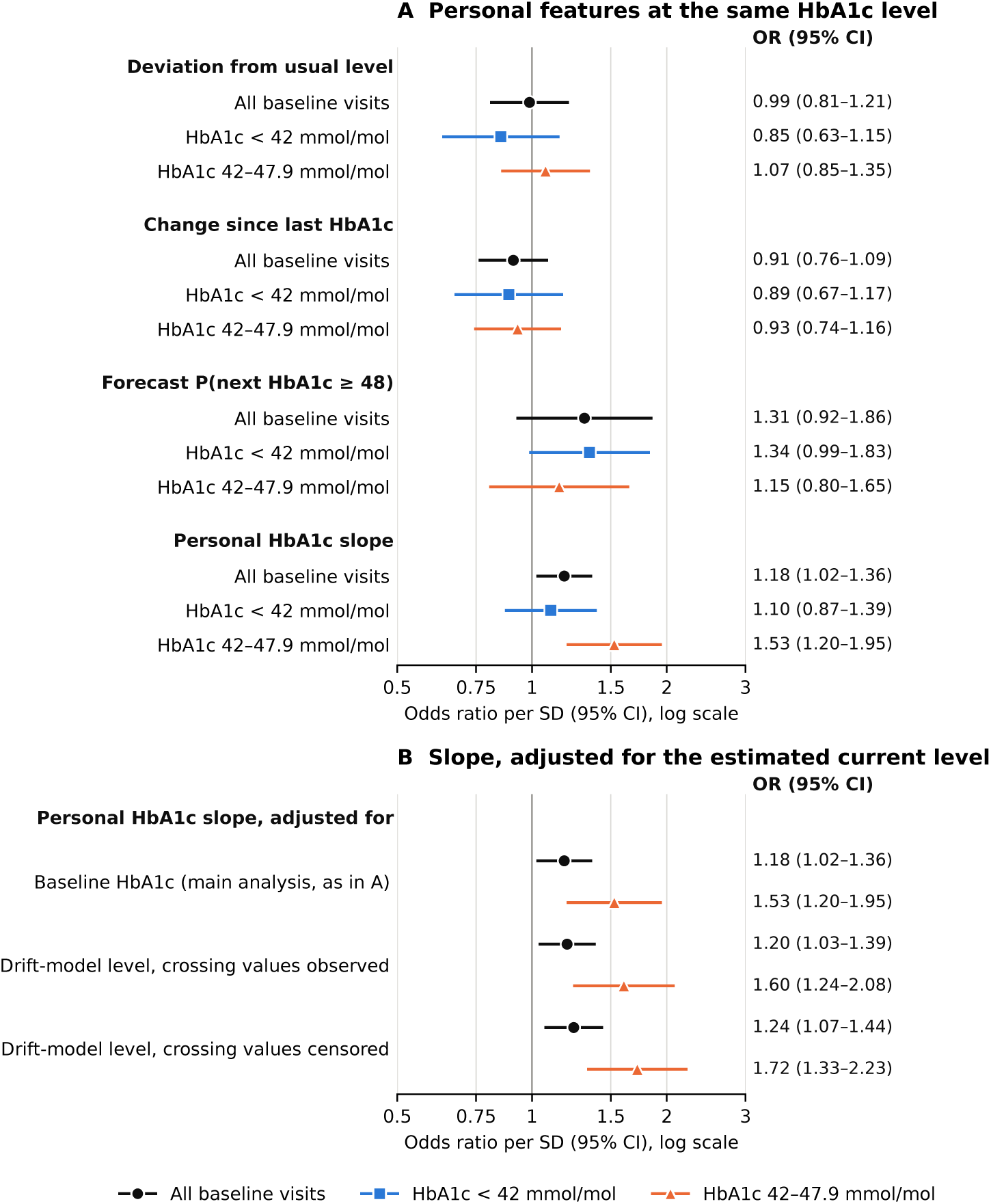
Adjusted odds ratios per SD with 95% CIs, shown on a logarithmic scale, from model (4). (A) Associations of the four personal-history features with incident diabetes, in all baseline visits and separately by baseline HbA1c range. (B) Personal-slope associations under three adjustments for current HbA1c: observed baseline HbA1c (repeated from panel A), the estimated underlying current level from the drift fit using observed threshold-crossing values, and the estimated level from the fit treating crossing values as censored at 48 mmol/mol; shown for all baseline visits and for HbA1c 42–47.9 mmol/mol. Black circles represent all baseline visits, blue squares HbA1c *<* 42 mmol/mol, and orange triangles HbA1c 42–47.9 mmol/mol. Each feature is standardised by its SD within the analysis group shown, so subgroup ORs refer to different absolute feature increments. Numerical results are provided in Supplementary Tables S7 and S10.

The slope association depended on the adjustment set: the OR was 0.85 (0.72–1.00) without adjustment and 1.05 (0.91–1.20) after adjustment for baseline HbA1c, and reached the fully adjusted estimate mainly through adjustment for age (Supplementary Table S8). The association therefore concerns a model-derived prognostic feature and does not by itself establish a persistent biological rate of change.

Separately fitted subgroup models gave ORs of 1.53 (1.20–1.95) for HbA1c 42–47.9 mmol/mol and 1.10 (0.87–1.39) below 42 mmol/mol, but these use subgroup-specific SDs and covariate effects; in a single interaction model there was no evidence that the association differed between the ranges (ratio of ORs, 42–47.9 versus *<* 42 mmol/mol, 1.03, 0.75–1.41).

The association remained when baseline HbA1c was replaced by the drift model’s estimate of the underlying current level (OR 1.20, 1.03–1.39; 1.24, 1.07–1.44 with the censored fit; Figure 4B). In simulated data in which risk depended only on the underlying level, so that the slope carried no information of its own, adjustment for the observed baseline value produced a spurious slope association (OR 1.26), which this adjustment removed (OR 1.02; Supplementary Section S4). The adjustment thus behaved as intended where the answer was known, and the persistence of the empirical association under the same adjustment is consistent with information in the slope beyond the current level.

Across the sensitivity analyses of slope construction, outcome definition, follow-up, missing covariates and deaths before follow-up, the slope OR ranged from 1.10 to 1.23 (Supplementary Tables S9 and S10). The lowest estimate (1.10, 0.96–1.26) arose when the baseline measurement was included in the slope while adjusting for observed baseline HbA1c, and rose to 1.17 (1.02– 1.35) when adjusting for the estimated current level instead. Among visits with at least two earlier measurements, the estimate was also insensitive to participation history (1.37–1.43; Supplementary Table S8).

### 3.4 Serial correlation and the observability of personal HbA1c slopes

The drift model (5) fitted better than the independent-noise model (1) in all three data sets: non-diabetic visits only, with threshold-crossing values added, and with these values censored (ΔAIC *−*140, *−*378 and *−*228; Table 4). Drift accounted for 77–81% of residual within-person variance.

How variation was divided between persistent slopes and drift, however, depended strongly on the crossing values (Table 4, Figure 5). With non-diabetic visits only, the time constant was poorly determined (50 years, 95% CI 12.5–87.6), and introducing drift reduced the estimated slope SD to 0.06, compared with 0.84 mmol/mol per decade under independent noise. Including the 401 crossing values (median 51 mmol/mol) shortened the time constant to 5.8 years (4.3–7.9) and gave a slope SD of 0.81 mmol/mol per decade; treating them as censored gave 26 years (14.4–43.2) and 0.33. Estimates changed smoothly with caps of 48 to 80 mmol/mol on the crossing values, with a larger change when the cap was removed, indicating sensitivity to the 16 values above 80 mmol/mol (Supplementary Table S11). In all drift fits, the intercept–slope correlation reached its boundary of 1. The data therefore did not determine robustly how variation was divided among personal intercept, persistent slope and slow drift, whereas allowing drift consistently improved fit, and its estimated share of residual variance was similar across fits.

**Figure 5:**
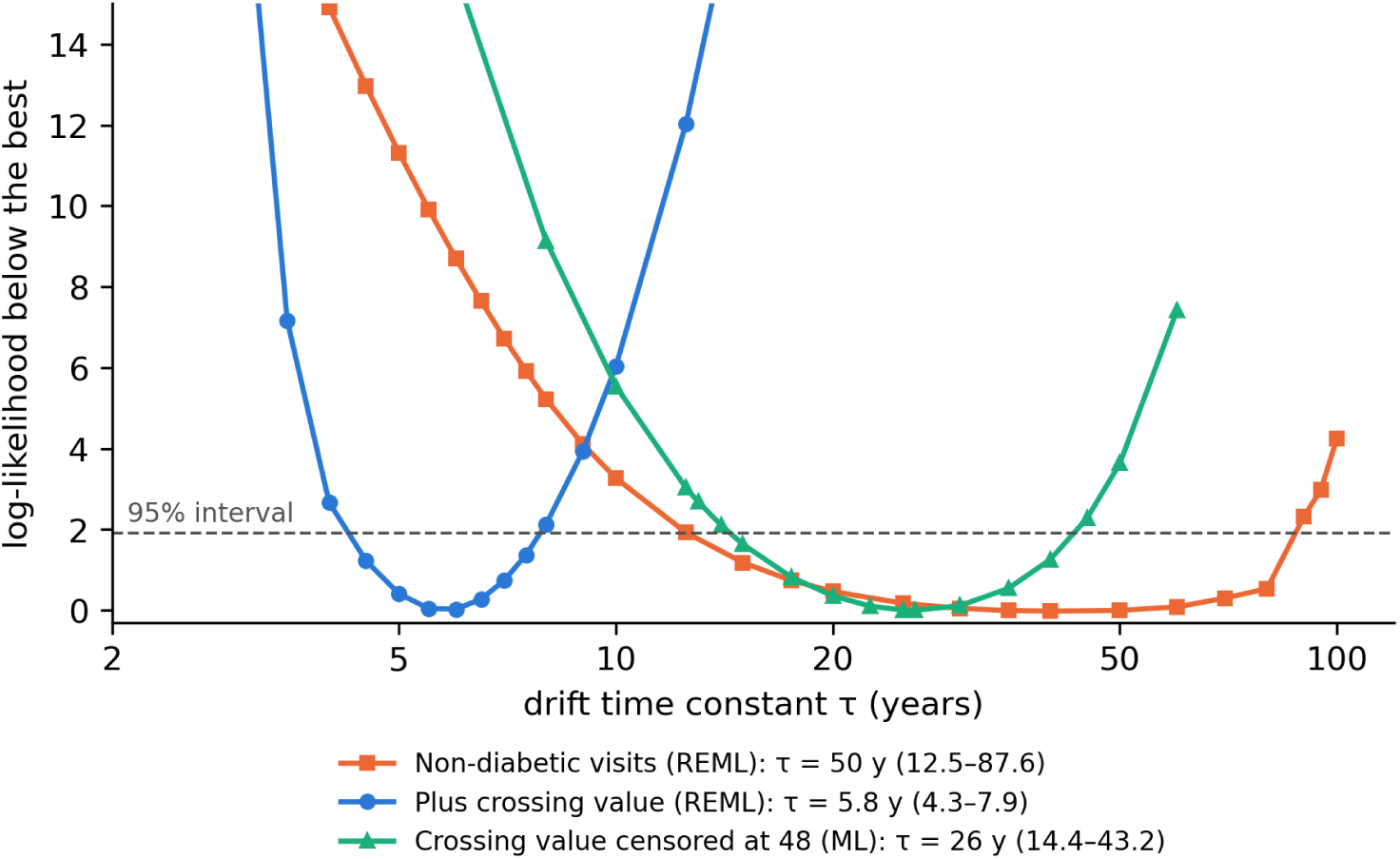
Profile likelihood for the drift time constant *τ* in the three data sets (non-diabetic visits only; plus threshold-crossing values; crossing values censored at 48 mmol/mol). For each fixed value of *τ*, the curve shows how much the log-likelihood falls below that of the best-fitting model; values of *τ* below the dashed line form the approximate 95% interval.

With the intercept–slope correlation set to zero, removing the assumed correlation between the personal intercept at age 65 and the slope (Section 2.9), persistent slopes were difficult to recover under the fitted models (Figure 6). With four-yearly measurements over up to 12 years, the expected correlation *r* between estimated and underlying slopes was at most 0.26 under the drift models (0.44 under independent noise). Quarterly measurements improved this little under drift, but raised it to 0.79 under independent noise; extending the observation period helped more, although after 30 years *r* was still 0.31–0.59 under drift. Under the fitted drift models and schedules examined, extending the observation period therefore improved slope recovery more than increasing measurement frequency. These calculations condition on the specified covariance parameters and do not incorporate their estimation uncertainty. At the fitted correlation of 1, *r* was already 0.69–0.75 with two measurements four years apart (Supplementary Table S12), because the slope is then a fixed multiple of the personal intercept at age 65; this reflects estimation of the intercept, not of change.

**Figure 6:**
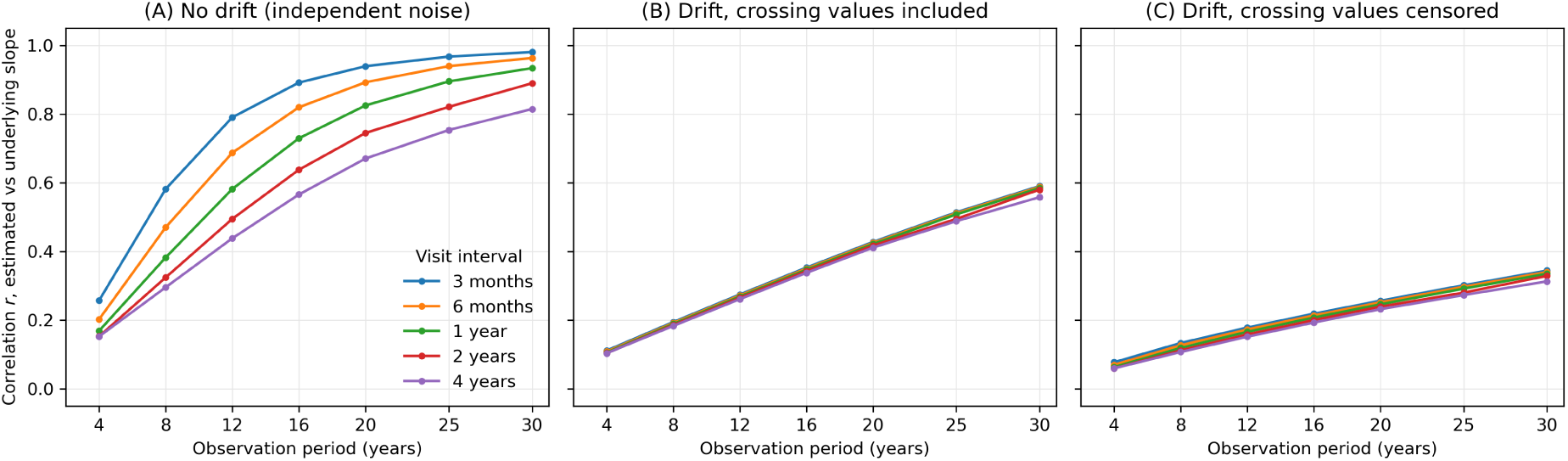
Observability of personal slopes: expected correlation *r* between the estimated and the underlying personal slope (equation (8)) for visit intervals of 3 months to 4 years and observation periods of 4 to 30 years. (A) Independent-noise model without drift. (B, C) Drift model fitted with the threshold-crossing values included (*τ* = 5.8 years) and censored at 48 mmol/mol (*τ* = 26 years). Under drift the curves for the five visit intervals nearly coincide: the length of the observation period, not the visit frequency, limits observability. Intercept–slope correlation set to zero (Section 2.9).

## 4 Discussion

In this work, we examined whether individual HbA1c trajectories of people without diabetes carry information about future diabetes beyond the current HbA1c value and conventional clinical risk factors. The question is motivated by the standard diagnostic approach, in which a single current value is compared with a fixed population threshold (World Health Organization, 2011; American Diabetes Association Professional Practice Committee, 2026). Our setting was a population-based cohort of middle-aged and older adults (ELSA) (Steptoe et al., 2013), in which HbA1c was measured at health visits several years apart over a long period.

As the unit of analysis, we used baseline visits: visits at which a participant was free of diabetes, had at least one earlier non-diabetic HbA1c value, and had HbA1c measured at the next visit (Section 2.2). Using a linear mixed model with independent residual noise (Laird and Ware, 1982; Verbeke and Molenberghs, 2000), we estimated the population trajectory and, by empirical Bayes (Robinson, 1991), each person’s expected trajectory from their earlier values (Section 2.3). From this model we derived the personal slope; the deviation of the baseline value from the person’s expected value, which measures how surprising the current value is given the person’s history; the change since the previous measurement; and a forecast, i.e. the predicted probability that HbA1c would reach the diabetic range at the next visit (Figure 1). Population parameters were estimated by cross-fitting (Chernozhukov et al., 2018), so that no person’s features were derived from a model fitted to their own data.

We added these features to a clinical prediction model including baseline HbA1c, age, sex and established clinical risk factors (Lindström and Tuomilehto, 2003; Wilson et al., 2007; Hippisley-Cox and Coupland, 2017; Noble et al., 2011; Abbasi et al., 2012). None of them meaningfully improved discrimination, in all baseline visits or in any subgroup examined, including visits with more than one earlier measurement (Section 3.2, Table 3, Supplementary Table S3).

To examine whether this reflected the choice of features rather than a lack of information in the histories, we also evaluated a time-aware Transformer designed for sparse, irregularly sampled data (Tipirneni and Reddy, 2022; Vaswani et al., 2017). It had access to the full observation histories and to additional variables in ELSA, such as waist circumference, further laboratory markers and self-rated health, and was pretrained to forecast future values (Li et al., 2020; Rasmy et al., 2021) (Section 2.7, Supplementary Table S5). The Transformer did not clearly improve on the clinical model, although a small improvement cannot be excluded. Pretraining improved forecasts of other variables but not of HbA1c, and did not improve discrimination; alternative configurations gave similar results (Supplementary Table S6). This agrees with previous findings that more flexible machine-learning methods do not necessarily outperform conventional regression models in clinical prediction (Christodoulou et al., 2019).

The absence of predictive improvement does not imply that the trajectories carry no relevant information. After adjustment for baseline HbA1c, age, sex and clinical risk factors, the personal slope was associated with incident diabetes (Section 3.3, Figure 4A), so the estimated slope carries information beyond the current HbA1c value. The association was clearest where the slope reflected observed within-person change, i.e. among visits with more than one earlier measurement (Supplementary Table S8). The deviation, the change since the previous measurement and the forecast showed no clear association (Supplementary Table S7).

Association and prediction are not in contradiction here. A predictor can be associated with an outcome while adding little to discrimination, particularly when established predictors such as baseline HbA1c already separate future cases well (Wald et al., 1999; Pepe et al., 2004; Ware, 2006; Cook, 2007). An association of the size observed for the slope would be expected to change the ranking of only few individuals once baseline HbA1c and clinical risk factors are known.

Two qualifications apply to the slope association itself. First, it depended on the adjustment set, particularly on adjustment for age; one possible contributor is that the estimated slope itself depends on the ages at which the earlier values were measured (Supplementary Section S3). Second, it concerns a model-derived prognostic feature and does not by itself establish a stable biological rate of change in each person.

The shortness of the histories may explain much of the gap between association and prediction. For most baseline visits only one earlier value was available (Section 3.1, Supplementary Table S2), so that the estimated slope was determined by how far that value lay from the population trajectory rather than by observed change. This may explain why the slope contributed little to prediction in most people and why the association was clearer when change had actually been observed. Imprecisely estimated slopes also attenuate associations with the underlying trajectories, analogous to regression dilution when exposures are measured with error (MacMahon et al., 1990; Clarke et al., 1999; Carroll et al., 2006).

The slope association was not readily explained by the earlier measurements merely providing a better estimate of the current HbA1c level. It remained when baseline HbA1c was replaced by the drift model’s estimate of the underlying current level (Figure 4B), whereas in simulated data in which risk depended only on the current level, the same adjustment largely removed the apparent slope association (Supplementary Section S4). The persistence of the empirical association is therefore consistent with information in the slope beyond the current level, although it remains conditional on the assumed drift model and cannot exclude this explanation entirely. The association was also similar across sensitivity analyses addressing slope construction, outcome definition, follow-up, missing covariates and deaths before follow-up (Supplementary Tables S9 and S10).

Deviations of HbA1c from a person’s own trajectory were not independent from visit to visit but persisted for years, as described by a serially correlated drift (Ornstein–Uhlenbeck) process (Uhlenbeck and Ornstein, 1930; Diggle, 1988; Taylor et al., 1994); this model fitted better than independent noise (Section 3.4, Table 4). How variation was divided between persistent slopes and drift, however, depended strongly on the treatment of the values at which HbA1c first crossed the diagnostic threshold, and in all drift fits the correlation between personal level and slope reached its boundary (Figure 5, Supplementary Table S11). The available data therefore could not reliably separate personal level, persistent slope and slow drift, whereas the presence of drift was consistent across specifications. Restricting trajectory analyses to disease-free observations can affect this separation (Hernán et al., 2004; Tobin et al., 2005), while measurements near disease onset may reflect the accelerating rise that often precedes diagnosis (Tabák et al., 2009; Heianza et al., 2012; Yu et al., 2023) and that neither persistent linear slopes nor stationary drift describe.

Serial correlation also limits what repeated measurements can reveal. When slopes could be inferred only from the measurements themselves, more frequent measurements improved slope recovery substantially under independent noise but little under drift, whereas longer observation periods helped more; even so, slopes remained poorly recovered over decades of observation (Section 2.9, Figure 6, Supplementary Table S12). Assuming independent variation would therefore overstate how reliably persistent individual change can be estimated, consistent with methodological work on the reliability of individual change (Willett, 1989; Raudenbush and Liu, 2001; Rast and Hofer, 2014; Brandmaier et al., 2018). The same applies to personal reference values: the index of individuality and the reference change value are usually interpreted with a single within-person variance (Harris, 1974; Fraser and Harris, 1989; Fraser, 2001; Coşkun et al., 2021), whereas under drift the expected difference between two measurements depends on the interval between them (equation (6)). Personal reference intervals remain a promising approach (Foy et al., 2025; Zhang et al., 2026; Shah et al., 2026), but their usefulness depends on the analyte, the outcome and how reliably a personal reference can be established. Much of the evidence on personal laboratory setpoints comes from electronic health records, in which testing frequency reflects clinical concern (Agniel et al., 2018; Goldstein et al., 2016; Sisk et al., 2021); scheduled measurements reduce this bias at the cost of sparse observations.

These results have implications for longitudinal biomarker monitoring. Estimating persistent individual slopes requires long histories, which favours long-term, routinely collected laboratory records, ideally in younger, working-age populations followed with regular health examinations. Detecting an emerging acceleration towards diabetes, by contrast, may benefit from more frequent measurements around the onset of progression and from models that allow the rate of change to increase, for example at a changepoint (Skates et al., 2001), or joint models of longitudinal markers and events (Tsiatis and Davidian, 2004; Rizopoulos, 2012; Diggle et al., 2015). The choice of observations also depends on the purpose of the model. When the outcome is crossing a diagnostic threshold, a person’s series ends at that point, so rising trajectories are cut off. A model that instead forecasts the biomarker itself could keep values above the threshold and exclude only measurements after the start of treatment, leaving the diagnostic decision to the clinician.

### 4.1 Strengths and limitations

An important strength of this work is the use of scheduled venous HbA1c measurements in a population cohort, which reduces bias associated with clinically triggered testing driven by symptoms or clinical suspicion. All representations of personal history were compared against a common clinical model, and personal features were constructed using participant-level cross-fitting, an approach that can be applied to other biomarkers. Comparing adjustment for observed and estimated underlying HbA1c helped distinguish trajectory information from better estimation of the current level. In addition, we extended the independent-noise model to account for serial correlation and quantitatively assessed how reliably individual slopes could be recovered under different measurement schedules.

The main limitation is the sparsity of the histories, with only one earlier value for most baseline visits. Conclusions about trajectories estimated from longer or denser histories are therefore limited, and the limited predictive gain from personal slopes may depend on the measurement intervals available in ELSA. The number of incident cases was modest (234), limiting the precision of performance and association estimates, particularly in subgroups. Incident diabetes was classified from a single HbA1c measurement at or above the diagnostic threshold, a reported diagnosis or medication use. The absence of confirmatory testing may have introduced outcome misclassification, including cases reflecting transient HbA1c elevations.

Another important limitation is the substantial loss to follow-up: more than 40% of eligible baseline visits had no HbA1c measurement at the next health visit. Inverse-probability weighting, the inclusion of interview-reported outcomes and scenarios for participants who died yielded similar slope associations (Supplementary Section S3), but selection bias from unmeasured factors remains possible. Furthermore, baseline visits with several earlier values came from participants who had survived and remained in the study for longer, so the stronger association in this subgroup may not apply to all participants.

Generalisability is also limited by the composition of the cohort. Almost all participants were of White ethnicity, limiting our ability to assess generalisability across ethnic groups. This is relevant because the relationship between HbA1c and glycaemia has been reported to differ between ethnic groups (Ziemer et al., 2010; Herman and Cohen, 2012). Generalisability to younger populations also remains uncertain.

A further limitation concerns the modelling of individual trajectories. The longitudinal features were derived from the independent-noise model and were not re-estimated under the better-fitting drift model. The latter’s parameters were not robustly determined: fits restricted to non-diabetic visits attributed almost no variation to persistent personal slopes, whereas variants allowing substantial persistent slopes required threshold-crossing measurements closely tied to the outcome (Table 4). The predictive value of features derived from alternative longitudinal models therefore remains uncertain. More generally, our representation of persistent change by linear personal slopes and of within-person fluctuation by independent noise or a stationary Ornstein– Uhlenbeck process may not capture nonlinear trajectories or other forms of within-person variation (Rasmussen and Williams, 2006; Hughes et al., 2017).

The observability calculations also depend on specific modelling assumptions. They used a reference scenario with the intercept–slope correlation set to zero, because a correlation of one would allow the slope to be inferred from the personal level rather than from observed change (Supplementary Section S4). These calculations therefore describe observability under the specified covariance assumptions rather than establishing a general limit on slope recovery.

Another limitation concerns the construction of features and the adjustment for the underlying current level. Participant-level cross-fitting excluded each participant’s own data when estimating the population trajectory parameters used to construct their features. However, these parameters were estimated using other participants’ measurements from all rounds, including rounds after a given baseline visit. Feature construction was therefore not strictly prospective. In addition, the drift model used to estimate the underlying current level was fitted to all participants, including the threshold-crossing values, rather than by cross-fitting. Performance under strictly prospective construction remains to be established.

Finally, the evaluation of the Transformer was limited to one main configuration and three variants without hyperparameter tuning. These results therefore do not establish the best performance achievable with this architecture.

## 5 Conclusion

ELSA provides an opportunity to investigate the value of personal laboratory histories in a population cohort with scheduled HbA1c measurements spanning almost two decades, reducing bias associated with clinically triggered testing. In this setting, estimated personal HbA1c slopes were associated with incident diabetes beyond baseline HbA1c and conventional clinical factors, most clearly where change had been directly observed, whereas deviations from estimated usual levels showed no clear association at the same baseline HbA1c. Nevertheless, neither these features nor a flexible sequence (Transformer) model showed clear evidence of improved discrimination beyond baseline HbA1c and conventional clinical factors, which captured nearly all the discrimination achieved by the evaluated models. Within-person variation was better described by serially correlated drift, as in an Ornstein–Uhlenbeck process, than by independent noise; under this description, persistent individual slopes were difficult to recover from measurements several years apart, which may explain part of this gap between association and prediction. These findings concern persistent individual slopes and do not exclude the predictive value of emerging nonlinear changes, including the acceleration in glycaemia that may precede diabetes.

More broadly, our results illustrate the importance of distinguishing longitudinal associations, incremental predictive value, and the observability of individual trajectories. This distinction is relevant beyond HbA1c: the usefulness of longitudinal biomarker information depends not only on its association with future disease, but also on whether that information can be recovered reliably from the available measurements.

## Supplementary material

The supplement follows the order of the analyses in the main text. Each section gives additional methods first, followed by the corresponding tables and figures.

### Supplementary Section S1: Cohort and trajectory model

Characteristics of the analysis set by baseline HbA1c range are given in Table S1, and the structure of the HbA1c histories before each baseline visit in Table S2. Figure S1 shows the distributional checks of the trajectory model (1) described in Section 2.3.

### Supplementary Section S2: Prediction analyses

This section provides details of the prediction analyses summarised in Section 2.4. It describes the sources and construction of the clinical predictors, specifies the final models A and B refitted to all baseline visits, with the equation needed to calculate predicted risks, reports differences in discrimination in subgroups of baseline visits, and describes the architecture, training and robustness of the time-aware Transformer.

#### Predictors

Body-mass index (BMI) was taken preferentially from the health-visit variable bmival. If it was missing, we used the measured BMI of the Harmonized ELSA file (mbmi); if this was also missing, BMI was calculated from the measured weight (mweight) and the most recent measured height (mheight) at or before that wave, retaining values between 12 and 80 kg/m^2^ (Gateway to Global Aging Data, 2023). Systolic blood pressure was ELSA’s derived variable sysval, the mean of the second and third of three seated readings. HDL cholesterol and triglycerides were measured by an external laboratory in the health-visit blood sample, fasting or non-fasting. Current smoking (smoken), vigorous physical activity at least once a week (vgactx_e) and blood-pressure medication (rxhibp) were self-reported at the interview of the same wave as the health visit and taken from the Harmonized ELSA file, in which participants without diagnosed hypertension are coded as not taking blood-pressure medication.

#### Final models

The final models in Table S4 were fitted to all 5,828 baseline visits using the same procedure as the fold-specific models. Predictors were transformed as shown, missing values were replaced by their medians on the transformed scale, and a missingness indicator was added for each covariate with missing values. All resulting columns, including the missingness indicators, were standardised using their means and SDs across all baseline visits after imputation. L2-penalised logistic regression maximised 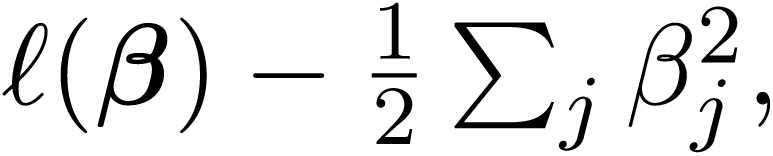 where *ℓ* is the log-likelihood summed over baseline visits and the penalty excludes the intercept (scikit-learn LogisticRegression, *C* = 1, lbfgs solver).

For a baseline visit, the predicted risk *p̂* is computed as

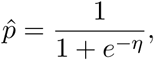

with

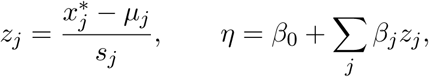

where *x_j_*\* denotes either a transformed and, if necessary, imputed predictor or a missingness indicator (0 if observed, 1 if missing). The corresponding imputation medians, means *µ_j_*, SDs *s_j_* and coefficients are given in Table S4. Because ln(years to next visit) is a predictor, *p̂* represents risk conditional on the specified interval to the next visit. The performance of these final models was not evaluated separately; reported performance refers to the out-of-fold predictions of the fold-specific models.

#### Time-aware Transformer

The model follows STraTS (Tipirneni and Reddy, 2022), building on attention architectures (Vaswani et al., 2017), earlier models for irregular clinical time series (Che et al., 2018; Horn et al., 2020; Shukla and Marlin, 2021) and pretrained EHR representations (Li et al., 2020; Rasmy et al., 2021). Each person’s history is a set of observations (time, variable, value); the network embeds every observation, lets the observations attend to each other, pools them into one vector, and adds static information. It is first trained to forecast future values (pretraining) and then to predict incident diabetes (fine-tuning). All settings are listed in Table S5.

Participants in the analysis cohort kept their folds from the main analysis, and the remaining pretraining participants were assigned to one of the five folds at random, so that no test-fold participant contributed to pretraining, fine-tuning, early stopping or normalisation. Pretraining used the complete observation histories of training-fold participants, including observations after their own baseline visits; this uses no information from test-fold participants but differs from a strictly chronological design.

### Supplementary Section S3: Association and sensitivity analyses

Odds ratios of the four features by HbA1c range are given in Table S7. The sensitivity analyses of the personal-slope association are grouped in three tables: adjustment set and measurement history (Table S8); follow-up, outcome definition and missing covariates (Table S9); and adjustment for the current level and construction of the slope (Table S10). Unless stated otherwise, all estimates in these tables are odds ratios per SD from model (4): adjusted for a natural cubic spline in baseline HbA1c, the clinical covariates of model B (age, sex, BMI, systolic blood pressure, HDL cholesterol, log triglycerides, smoking, weekly vigorous activity, blood-pressure medication) and ln(years to next visit), with median imputation and missingness indicators and participant-clustered standard errors; the slope is standardised by its SD within the analysed set of baseline visits. Counts of baseline visits and cases refer to all baseline visits in the analysed set.

#### Adjustment set

Without adjustment, the slope OR was below one. This may partly reflect the dependence of the estimated slope on the person’s HbA1c level through the fitted random-effects covariance and empirical Bayes shrinkage (estimated slopes were lower at higher HbA1c; Table 1, footnote d; Supplementary Table S1), together with the strong association between baseline HbA1c and diabetes risk. Estimated in separate fully adjusted models, the slope OR was 0.98 (0.80–1.20) with one earlier value, where the estimated slope correlates strongly with the person’s level (*r* = *−*0.44 with baseline HbA1c), and 1.39 (1.11–1.74) with at least two; these separate estimates differ partly because the covariate effects and the SD used for scaling are estimated within each subgroup, and the interaction model gave no evidence of a difference. To examine the role of the covariates in the slope association, the clinical covariates were added one at a time to a model adjusted only for the HbA1c spline and follow-up duration, and removed one at a time from the full model (Table S8). Only age changed the estimate materially. The estimated slope had little residual linear association with age after adjustment for the HbA1c spline and follow-up duration (partial correlation 0.01 overall; 0.02 with one and *−*0.01 with at least two earlier measurements); these correlations do not, however, establish why adjustment for age changed the slope OR. One relevant property of the estimator is that its dependence on an earlier HbA1c value varies with the age at measurement. With a single earlier value *y*_1_ at *a*_1_ = (age_1_ *−* 65)*/*10 and deviation 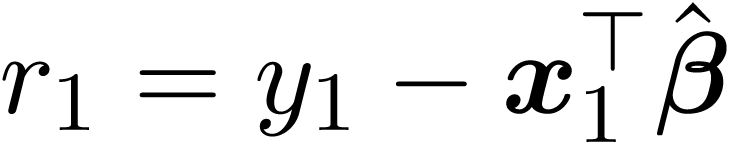 from the population trajectory, the estimated slope deviation is

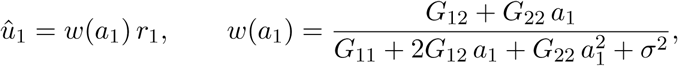

When *G*_12_ = 0, *w*(*a*_1_) changes sign at age 65: a positive deviation from the population trajectory lowers the estimated slope when measured before age 65 and raises it when measured after, and a negative deviation has the opposite effect; in general, the sign changes at age 65 *−* 10 *G*_12_*/G*_22_, which with the rounded parameters of Table 2 is about 66 years (illustrative, as the parameters differ slightly between folds). Even without observed within-person change, the estimated slope therefore depends on both the deviation of the earlier measurement and the age at which it was obtained. This age dependence of the feature is one plausible contributor to the effect of age adjustment, but the calculation does not by itself establish the mechanism.

#### Measurement history

Among baseline visits with at least two earlier HbA1c measurements, the slope association was additionally adjusted, one at a time, for a natural cubic spline in age (replacing the linear term), for birth year, and for the time since the first HbA1c measurement, and was estimated separately below and above age 70; strata with fewer than 10 cases were not estimated.

#### Loss to follow-up

For inverse-probability weighting, the probability of having HbA1c at the next health visit was estimated among all eligible baseline visits by a ridge-penalised logistic regression (as in the prediction models) on age, sex, HbA1c, BMI, systolic blood pressure, HDL cholesterol, log triglycerides, smoking, weekly vigorous activity, blood-pressure medication, education, employment, the number of earlier HbA1c measurements and the baseline round (AUC 0.69). Predicted probabilities were bounded below at 0.05 before calculating inverse-probability weights, which were then scaled to a mean of 1 in the analysis set. The maximum weight was 4.75 (99th percentile 2.23), and the effective sample size was 5,259 of 5,828 baseline visits (90%). The weighted analysis assumes that follow-up is independent of the missing outcome given these variables.

For the extended outcome set, baseline visits without HbA1c at the next health visit were added if the participant answered the diabetes question at an interview in the next health-visit round, or reported diabetes at any interview between the baseline visit and the next round. A report of no diabetes before the next round did not qualify, because diabetes could still have developed by that round. Incident diabetes in these visits was defined by the interview reports, or by fasting glucose at a next visit without valid HbA1c. Because known cases could enter more readily than non-cases, inclusion depended partly on the outcome. This analysis is therefore a sensitivity check rather than a removal of selection bias; its interpretation depends on the inclusion mechanism, including whether inclusion varied with the covariates, and on differences between interview-based and laboratory-based ascertainment of diabetes.

Deaths were identified from the harmonised interview status and the wave-11 end-of-life file. For participants who died before the next health visit, hypothetical diabetes outcomes were drawn with probabilities predicted by a logistic model fitted to the analysis set, evaluated at the person’s values at the baseline visit, with the follow-up interval set to the median of the baseline round in both outcome generation and the subsequent association model. The coefficients of the risk model were held fixed, so the pooled intervals do not reflect uncertainty in that model. The predicted probabilities were multiplied by 1, 2 or 3 and capped at 1; the multiplier acts on the probability, not on the odds. For each scenario, 50 sets of outcomes were generated, the association model was refitted on all baseline visits, and estimates were pooled with Rubin’s rules. In the first set of scenarios, the outcome-generating model included the personal slope; in the second, it did not. Because the slope is correlated with baseline HbA1c and the other predictors, the hypothetical outcomes in these scenarios are not independent of the slope, but the outcome-generating model contains no explicit slope term. Death is a competing event, and the hypothetical outcomes describe a next visit that did not take place, not diabetes incidence after death.

#### Outcome definition

For the outcome defined by HbA1c alone, incident diabetes was HbA1c *≥* 48 mmol/mol at the next health visit. Visits that were cases only through fasting glucose, a reported diagnosis or medication were either excluded or classified as non-cases (Table S9); the first version selects visits using follow-up information and is therefore a sensitivity analysis in a selected sample. Because no case occurred at the lowest baseline HbA1c values, the spline was evaluated at max(HbA1c*, c*), with *c* the lowest baseline HbA1c of a case in each analysed sample, and the missingness indicators for HDL cholesterol and triglycerides were replaced by a single indicator that either value was missing.

#### Missing covariates

For multiple imputation, the seven clinical covariates with missing values (BMI, systolic blood pressure, HDL cholesterol, log triglycerides, smoking, weekly vigorous activity and blood-pressure medication) were imputed 20 times by chained equations (scikit-learn IterativeImputer with Bayesian ridge regression and draws from the posterior predictive distribution, 10 iterations per data set); imputed values of binary covariates were clipped to [0, 1] and rounded to 0 or 1. The imputation model included the outcome, the four personal features, baseline HbA1c, age, sex, ln(years to next visit) and the health-visit round. Imputation was performed at the level of baseline visits and did not model the dependence between visits of the same participant. This sensitivity analysis therefore used an approximate imputation model: binary variables were imputed with continuous models, and within-participant dependence was not represented. In each imputed data set, the association model was fitted without missingness indicators and with participant-clustered standard errors; the estimates and these variances were pooled with Rubin’s rules, with Rubin’s degrees of freedom for the *t* reference distribution (Rubin, 1987; White et al., 2011; van Buuren, 2018). The analysis was run for all four features in all baseline visits and in the HbA1c range 42–47.9 mmol/mol, and for the personal slope among baseline visits with at least two earlier HbA1c measurements.

### Supplementary Section S4: Drift model and slope observability

#### Fit of the trajectory models

Figure S2 compares the empirical correlation of residuals between pairs of health-visit rounds with the correlations implied by the independent-noise and drift models, and Figure S3 the empirical semivariogram with the curves implied by the fitted models (equation (6)). Both use deviations from the population trajectory, *d_ij_* = *y_ij_ −* ***x***^T^***β̂***, with the fixed effects of the independent-noise model; these deviations retain the personal intercept and slope. Model-implied correlations between two rounds were approximated by evaluating the fitted model at the mean ages of the participants measured in both rounds. The model curves describe the unconditional covariance structure; because the observed series were restricted by diabetes status and threshold crossing, the empirical correlations and semivariograms need not coincide exactly with these curves, even under the assumed model.

The profile likelihood for *τ* was evaluated at fixed values between 0.5 and 100 years (in steps of 0.5 years between 3 and 8 years, more widely spaced elsewhere, with additional values where the profile was flat near the interval limits). The approximate 95% interval comprised the values of *τ* whose log-likelihood was within 1.92 (half the 95th percentile of a *χ*^2^ distribution) of the maximum, with limits interpolated on log *τ* between grid points; because *τ* is weakly identified, these intervals are approximate. The drift and independent-noise models have the same fixed-effect specification. Likelihood-ratio statistics and ΔAIC compared models fitted to the same observations with the same likelihood convention: REML for the uncensored data sets and maximum likelihood for the censored fit, in which the independent-noise model was obtained by setting the drift variance to zero. Because *σ_w_* = 0 lies on the boundary of the parameter space and *τ* is undefined under the independent-noise model, the likelihood-ratio statistic does not follow the usual *χ*^2^ distribution (Self and Liang, 1987); we report it descriptively, without a *p*-value.

#### Threshold-crossing values

Series ending because of other diabetes criteria (fasting glucose, self-reported diagnosis or medication) with HbA1c below 48 mmol/mol were not extended by a crossing value. Including the crossing value makes the HbA1c-based stopping rule depend on an observed rather than an excluded value; under a correctly specified model, and if other dropout is missing at random, likelihood-based estimation then requires no separate model of the dropout process (Rubin, 1976; Diggle and Kenward, 1994; Little and Rubin, 2019). This does not address selection arising from diagnosis by other criteria, treatment or death.

In the censored fit, a participant with a crossing value contributed the density of their non-diabetic values multiplied by the model probability that the next value was *≥* 48 mmol/mol given those values; all other participants contributed the density of their values as before. The censored model was fitted by maximum likelihood; for the uncensored fits, maximum likelihood and REML gave nearly identical estimates. We verified the censored likelihood against an independent implementation based on full-covariance Gaussian conditioning. To examine how strongly the uncensored fit depended on the largest crossing values, we also refitted it with the crossing values capped at 48 to 80 mmol/mol (Table S11, Figure S4).

#### Simulation of the current-level adjustment

The simulation used to check the level adjustment (Section 3.3) generated 12,000 hypothetical participants with two to four HbA1c values about four years apart, the first at age 55–70, under a drift model with parameters close to the fit including crossing values (intercept SD 3.0 mmol/mol, slope SD 0.8 mmol/mol per decade, intercept–slope correlation 1, drift SD 2.8 mmol/mol, *τ* = 5.6 years, independent SD 1.3 mmol/mol). The population trajectory was 38.6 + 0.94 *a* mmol/mol, with *a* = (age *−* 65)*/*10 and no sex effect, and the spacing between values varied with SD 0.4 years (function simulate_selftest in the analysis code). The last measurement defined baseline. Histories containing any HbA1c value *≥* 48 mmol/mol were excluded, leaving 11,399 participants. A subsequent binary diabetes outcome was generated with probability expit*{−*3.2 + 0.45(*L −* 40)*}*, where *L* was the underlying level at baseline, yielding 769 cases; the personal slope therefore carried no information beyond that level. The slope feature was the empirical Bayes slope from the earlier values under an independent-noise model with fixed parameters (intercept SD 3.4 mmol/mol, slope SD 1.1 mmol/mol per decade, correlation 0.69, residual SD 2.6 mmol/mol); the underlying level was estimated with the true drift parameters, which were therefore known rather than re-estimated. The slope association was estimated once, in a single simulated data set, with a natural cubic spline (4 df) in either the observed last value or the estimated level and no other covariates, and with the slope standardised by its SD within each analysed group: OR 1.26 (95% CI 1.12–1.42) and 1.02 (0.90–1.16) in all simulated participants, and 1.24 (1.11–1.38) and 1.09 (0.97–1.23) among those with a last value of 42–47.9 mmol/mol. An intercept–slope correlation of 1 was chosen to reflect the boundary estimates of the fitted drift models. The simulation illustrates that adjustment for the estimated current level can substantially attenuate an apparent slope association under this particular scenario; it does not establish that the empirical association is free of this mechanism.

#### Observability

Table S12 gives the expected correlation between estimated and underlying slopes with the intercept–slope correlation set to zero, as in the reference observability calculation (Section 2.9), and at its fitted value. Measurements were equally spaced and, where the history length was a multiple of the interval, the last was at age 65, the reference age of the model; a four-year history therefore contains two four-yearly values. The four-yearly 30-year schedule comprised values at ages 35 to 63 (28-year span), whereas the quarterly schedule spanned the full 30 years, so this comparison differs in duration as well as frequency. Calculations condition on the specified covariance parameters and do not incorporate their estimation uncertainty. The schedules describe what repeated measurements over a given span can reveal; they do not reproduce the primary slope estimator, which used only values before the baseline visit and, for 61% of baseline visits, a single earlier value.

### Supplementary Section S5: Reporting checklists

The STROBE and TRIPOD+AI checklists are given in Tables S13 and S14.

## Declarations

### Ethics

Secondary analysis of de-identified data obtained under the UK Data Service End User Licence; no additional ethical approval was required. ELSA received ethical approval from the relevant NHS Research Ethics Committees, and participants gave informed consent.

### Protocol and registration

No protocol was prepared in advance and the study was not registered.

### Patient and public involvement

Patients and the public were not involved in the design, conduct or reporting of this study.

### Data availability

ELSA data are available from the UK Data Service (SN 5050) under an End User License (https://doi.org/10.5255/UKDA-SN-5050-37).

### Code availability

The analysis code is available at https://github.com/dapias/ elsa-hba1c-trajectories (release v1.0.0).

### Use of AI-assisted tools

Claude by Anthropic was used during manuscript preparation to support language editing, clarity, grammar, and phrasing. The authors reviewed and verified all artificial intelligence–assisted text and remain fully responsible for the accuracy, originality, and integrity of the manuscript.

## Data Availability

ELSA data are available from the UK Data Service (SN 5050) under an End User Licence: https://doi.org/10.5255/UKDA-SN-5050-37. Under the terms of this licence, the authors cannot share the individual-level data directly. The analysis code is publicly available at https://github.com/dapias/elsa-hba1c-trajectories (release v1.0.0). Aggregate results are contained in the manuscript and its supplementary material.

https://github.com/dapias/elsa-hba1c-trajectories

## Acknowledgements

We thank Saadet Melisa Kemiksiz for her support in reviewing the ELSA documentation and with the literature review. ELSA is funded by the National Institute on Aging (R01AG017644) and by UK Government Departments coordinated by the National Institute for Health and Care Research (NIHR). The data were made available through the UK Data Service. Neither the original collectors of the data nor the UK Data Service bear any responsibility for the analyses or interpretations presented here.

---

**Figure S1:**
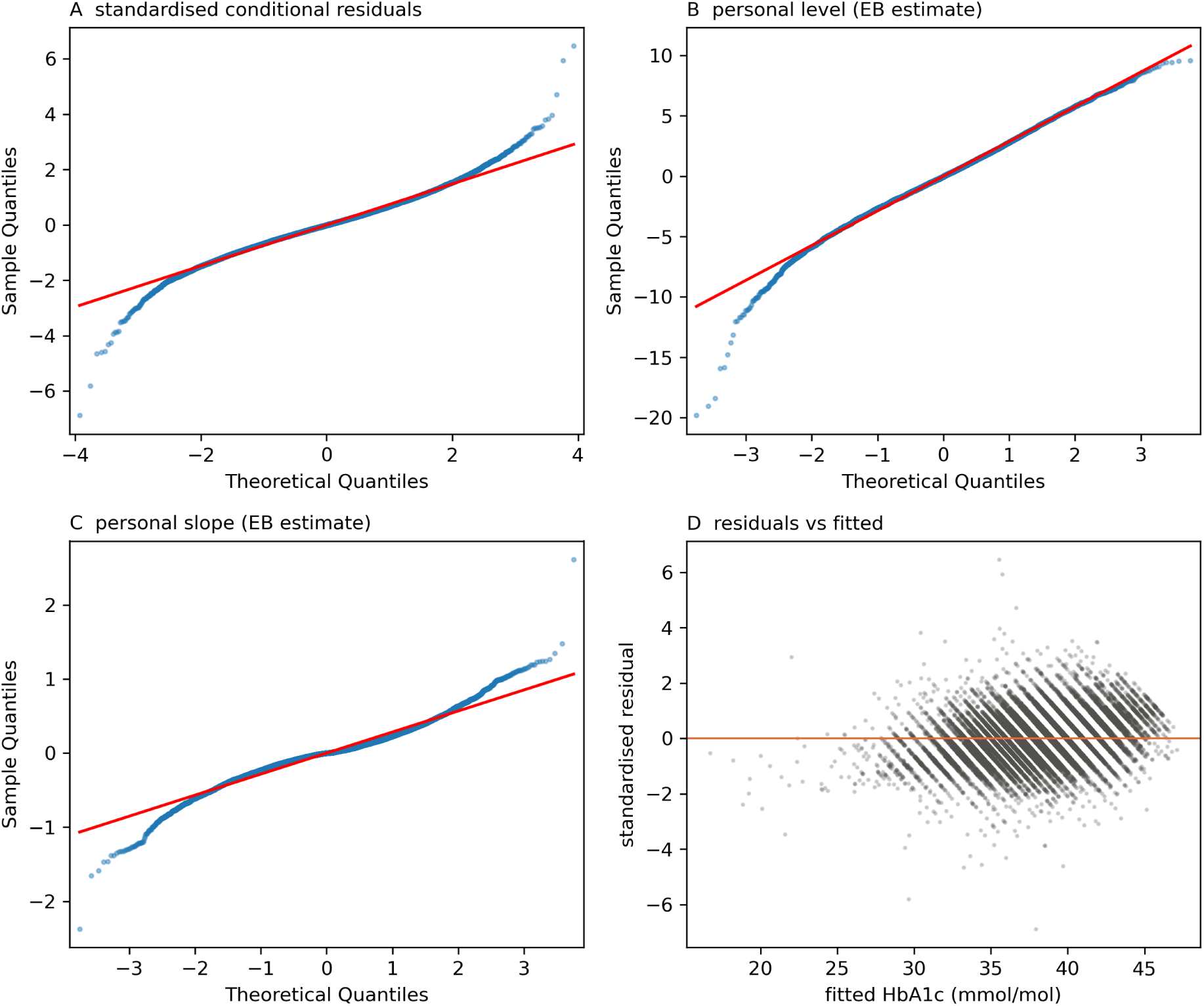
Diagnostics of the trajectory model (1) fitted to all non-diabetic visits. Normal quantile– quantile plots of (A) the standardised conditional residuals, (B) the empirical Bayes personal levels at age 65 and (C) the empirical Bayes personal slopes; (D) standardised conditional residuals against fitted values. Because empirical Bayes estimates are shrunk towards the population, panels B and C give only an approximate check of the assumed normality of the random effects.

**Figure S2:**
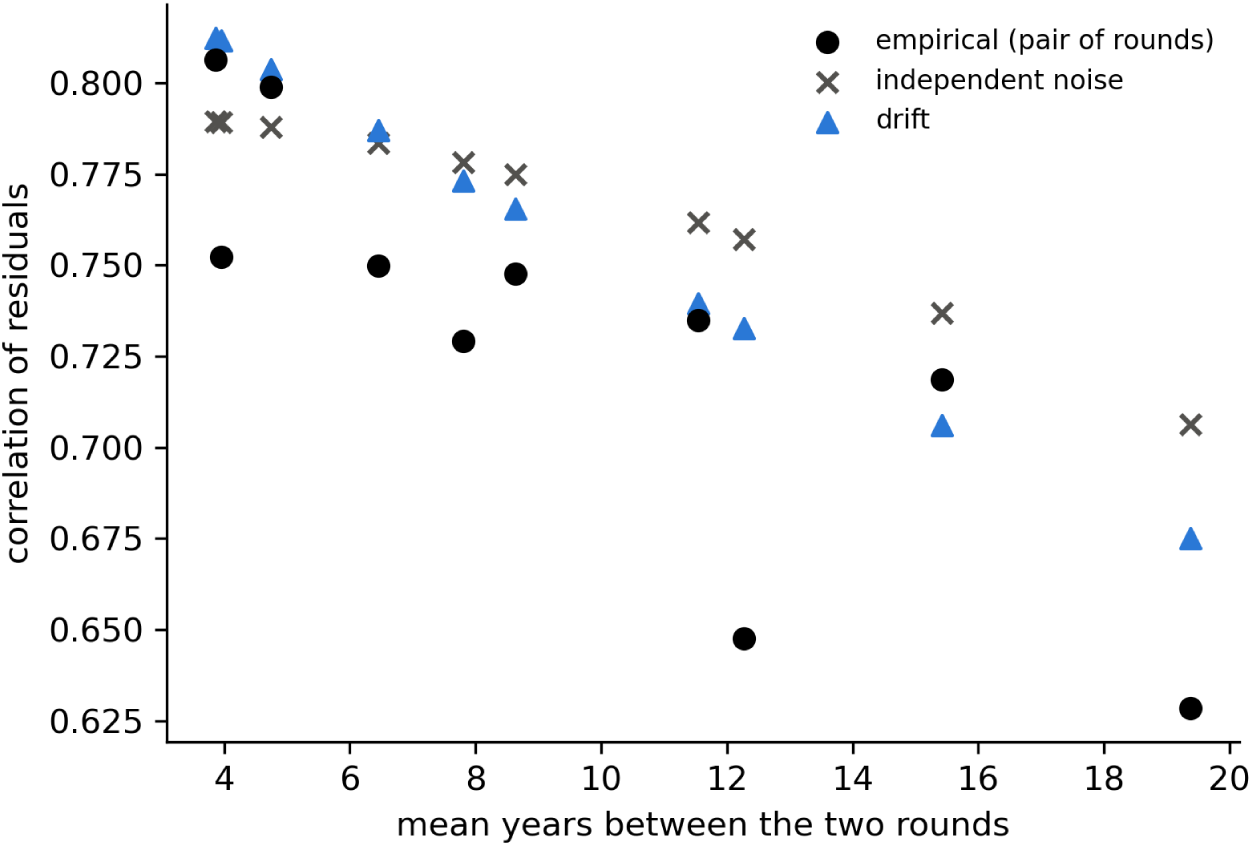
Correlation of residuals between pairs of health-visit rounds against the mean time between them: empirical and implied by the independent-noise and drift models (non-diabetic visits).

**Figure S3:**
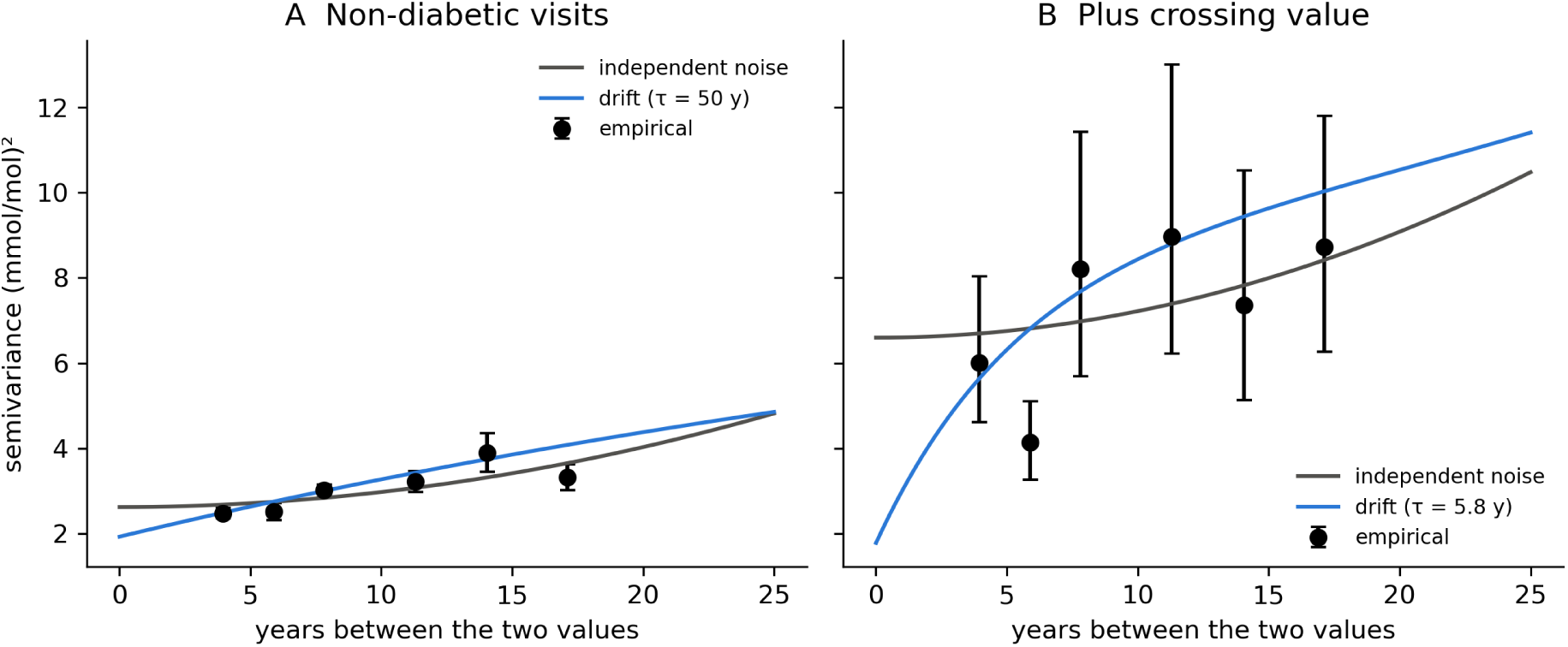
Empirical semivariogram of the residuals from the population trajectory (points, 95% participant-cluster bootstrap intervals) and the curves implied by the fitted independent-noise and drift models (equation (6)), for (A) non-diabetic visits only and (B) with the threshold-crossing values added. No empirical semivariogram is shown for the censored specification, because the crossing magnitudes are deliberately omitted from that analysis. The scheduled health-visit rounds provide observations at a limited number of lag ranges.

**Figure S4:**
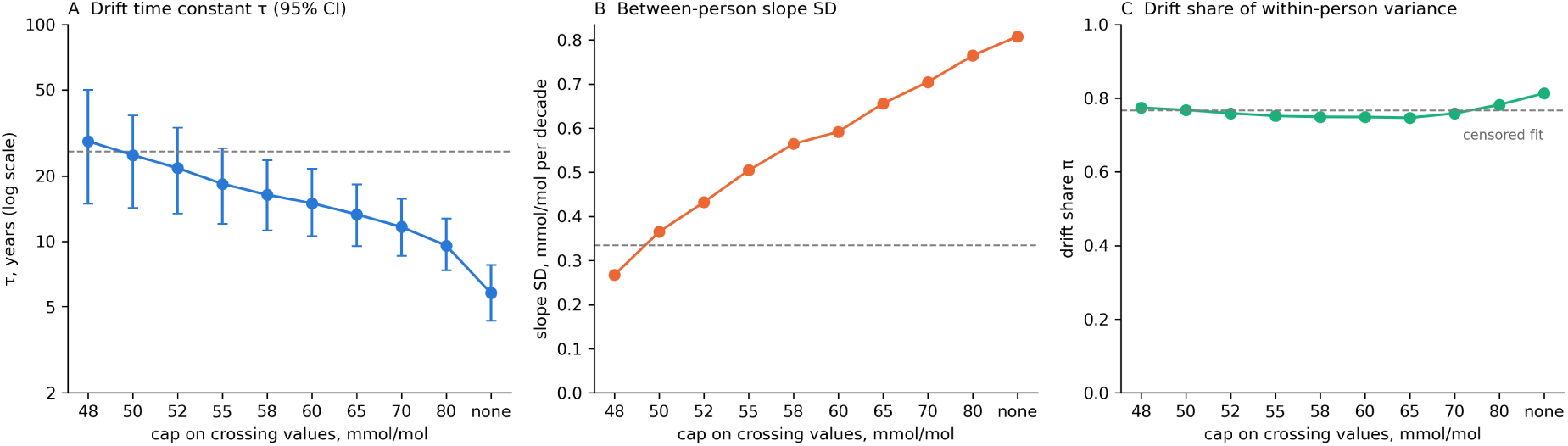
Drift model by cap on the threshold-crossing values: (A) drift time constant *τ* (log scale, 95% CI), (B) between-person slope SD and (C) drift share *π*. Dotted line: 55 mmol/mol, the upper quartile of the crossing values (Table S11).

**Table S1:** Characteristics at the baseline visit by HbA1c range (analysis set). Baseline visits with HbA1c 48 mmol/mol are excluded by design (diabetes at baseline). Personal slopes are estimated from the earlier values only. Percentages of categorical covariates refer to visits with non-missing values.

|  | HbA1c < 42 mmol/mol | HbA1c 42–47.9 mmol/mol |
|---|---|---|
| Baseline visits, n | 4,634 | 1,194 |
| Persons, n | 2,997 | 896 |
| Incident diabetes, n (%) | 81 (1.7) | 153 (12.8) |
| Age, years, mean (SD) | 67.4 (7.2) | 68.9 (7.6) |
| Women, n (%) | 2,618 (56.5) | 703 (58.9) |
| HbA1c, mmol/mol, mean (SD) | 37.5 (2.7) | 43.5 (1.5) |
| HbA1c, mmol/mol, median (IQR) | 38.0 (36.0–39.9) | 43.0 (42.1–44.3) |
| HbA1c, %, mean (SD) | 5.58 (0.25) | 6.13 (0.14) |
| Personal slope, mmol/mol per decade, mean (SD) | 0.96 (0.16) | 0.89 (0.18) |
| BMI, kg/m<sup>2</sup>, mean (SD) | 27.3 (4.5) | 28.6 (5.0) |
| Waist, cm, mean (SD)<sup>a</sup> | 94.3 (12.3) | 97.4 (12.9) |
| Systolic BP, mmHg, mean (SD) | 131.8 (16.8) | 133.5 (17.6) |
| BP medication, n (%) | 1,216 (26.5) | 411 (34.7) |
| HDL cholesterol, mmol/l, mean (SD) | 1.68 (0.46) | 1.58 (0.42) |
| Triglycerides, mmol/l, median (IQR) | 1.2 (0.9–1.7) | 1.4 (1.0–2.0) |
| Current smoker, n (%) | 354 (7.7) | 152 (12.8) |
| Vigorous activity weekly, n (%) | 1,828 (39.5) | 385 (32.2) |
| Fasting blood sample, n (%) | 3,475 (75.0) | 827 (69.3) |
| *Earlier HbA1c values, n (%)* |  |  |
| 1 | 2,829 (61.0) | 723 (60.6) |
| 2 | 1,420 (30.6) | 395 (33.1) |
| 3 | 385 (8.3) | 76 (6.4) |
| Years to next visit, median (IQR) | 4.2 (3.9–6.1) | 4.1 (3.8–5.6) |
| *Missing, %* |  |  |
| BMI; waist | 1.2; 27.2 | 1.6; 16.6 |
| Systolic BP; BP medication | 4.0; 1.1 | 5.4; 0.8 |
| HDL cholesterol; triglycerides | 0.7; 0.6 | 0.6; 0.5 |
| Smoking; physical activity | 0.3; 0.0 | 0.3; 0.0 |

<sup>a</sup> Waist circumference was not available in the round 8/9 health-visit file.

**Table S2:**
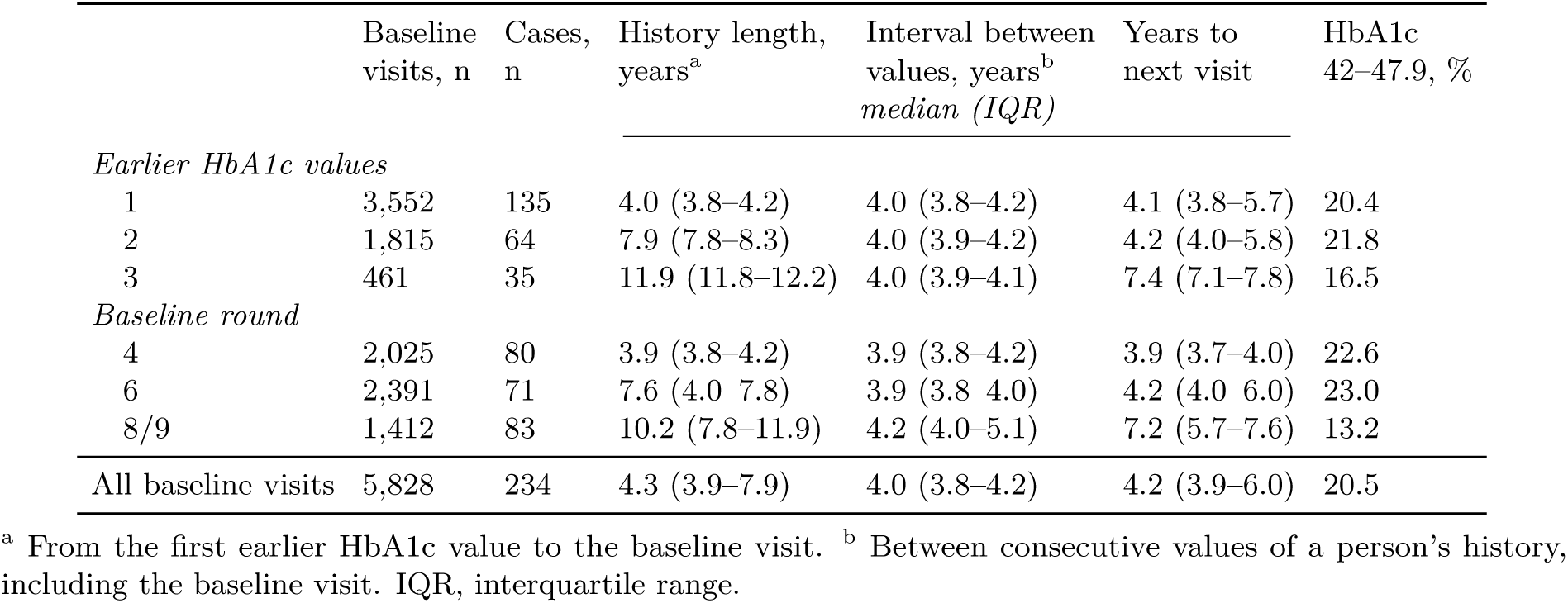
Structure of the HbA1c measurement history before each baseline visit (analysis set). Rounds 8 and 9 form one health-visit round.

|  | Baseline visits, n | Cases, n | History length, years<sup>a</sup> | Interval between values, years<sup>b</sup> | Years to next visit | HbA1c 42–47.9, % |
|---|---|---|---|---|---|---|
|  |  |  | *median (IQR)* |  |  |  |
| *Earlier HbA1c values* |  |  |  |  |  |  |
| 1 | 3,552 | 135 | 4.0 (3.8–4.2) | 4.0 (3.8–4.2) | 4.1 (3.8–5.7) | 20.4 |
| 2 | 1,815 | 64 | 7.9 (7.8–8.3) | 4.0 (3.9–4.2) | 4.2 (4.0–5.8) | 21.8 |
| 3 | 461 | 35 | 11.9 (11.8–12.2) | 4.0 (3.9–4.1) | 7.4 (7.1–7.8) | 16.5 |
| *Baseline round* |  |  |  |  |  |  |
| 4 | 2,025 | 80 | 3.9 (3.8–4.2) | 3.9 (3.8–4.2) | 3.9 (3.7–4.0) | 22.6 |
| 6 | 2,391 | 71 | 7.6 (4.0–7.8) | 3.9 (3.8–4.0) | 4.2 (4.0–6.0) | 23.0 |
| 8/9 | 1,412 | 83 | 10.2 (7.8–11.9) | 4.2 (4.0–5.1) | 7.2 (5.7–7.6) | 13.2 |
| All baseline visits | 5,828 | 234 | 4.3 (3.9–7.9) | 4.0 (3.8–4.2) | 4.2 (3.9–6.0) | 20.5 |

<sup>a</sup> From the first earlier HbA1c value to the baseline visit. <sup>b</sup> Between consecutive values of a person’s history, including the baseline visit. IQR, interquartile range.

**Table S3:** Differences in out-of-fold AUC between prediction models within subgroups of baseline visits. Subgroup analyses used the existing predictions without refitting the models. Differences were calculated using the same baseline visits for both models in each bootstrap replicate. The 95% CIs were obtained from a participant-cluster bootstrap with 1,000 replicates, conditional on the fitted predictions and without refitting the models. Results for all baseline visits are given in Table 3.

| Visits (cases) | HbA1c < 42 mmol/mol<br>4,634 (81) | 1 earlier value<br>3,552 (135) | ≥ 2 earlier values<br>2,276 (99) | Rounds 4 and 6<sup>a</sup><br>4,416 (151) |
|---|---|---|---|---|
| B − A | +0.049 (0.020 to 0.080) | +0.019 (−0.001 to 0.039) | +0.016 (−0.002 to 0.033) | +0.023 (0.005 to 0.040) |
| C − B | +0.001 (−0.003 to 0.005) | +0.001 (−0.001 to 0.003) | 0.000 (−0.002 to 0.002) | +0.001 (−0.001 to 0.003) |
| D − B | −0.006 (−0.015 to 0.003) | −0.002 (−0.006 to 0.002) | −0.001 (−0.006 to 0.004) | 0.000 (−0.003 to 0.004) |
| E − B | +0.004 (−0.007 to 0.015) | +0.002 (−0.005 to 0.008) | −0.003 (−0.009 to 0.004) | 0.000 (−0.006 to 0.006) |

Models as in Table 3: A, HbA1c, age and sex; B, A plus clinical factors; C, B plus change since the last value; D, B plus deviation *z* and personal slope; E, B plus logit of forecast probability.
<sup>a</sup> Baseline visits with an interval of about four years to the next health visit.

**Table S4:** Final models A and B, refitted to all 5,828 baseline visits using L2-penalised logistic regression on standardised predictors (Supplementary Section S2). These are distinct from the five fold-specific models used to generate the out-of-fold predictions in Table 3. Imputation medians were calculated from observed values on the transformed scale; means and SDs were calculated after imputation and before standardisation. Model A and Model B columns report coefficients for standardised predictors; intercepts are unpenalised. One missingness-indicator term is withheld (footnote a); its contribution to the log-odds is less than 0.001 in absolute magnitude when vigorous activity is observed. The complete specification can be regenerated by running the analysis code on the licensed data.

| Predictor | Transformation | Median | Mean | SD | Model A | Model B |
|---|---|---|---|---|---|---|
| Intercept | – | – | – | – | −4.053 | −4.197 |
| HbA1c, mmol/mol | none | – | 38.74 | 3.47 | 1.469 | 1.341 |
| Age, years | none | – | 67.71 | 7.30 | −0.147 | −0.142 |
| Female sex | 0/1 | – | 0.57 | 0.50 | −0.067 | −0.038 |
| Years to next visit | ln | – | 1.56 | 0.26 | 0.507 | 0.531 |
| BMI, kg/m<sup>2</sup> | none | 26.97 | 27.58 | 4.62 | – | 0.310 |
| Systolic BP, mmHg | none | 131.00 | 132.06 | 16.61 | – | 0.141 |
| HDL cholesterol, mmol/l | none | 1.60 | 1.66 | 0.45 | – | −0.109 |
| Triglycerides, mmol/l | ln | 0.26 | 0.28 | 0.47 | – | 0.204 |
| Current smoker | 0/1 | 0.00 | 0.09 | 0.28 | – | −0.024 |
| Weekly vigorous activity | 0/1 | 0.00 | 0.38 | 0.49 | – | −0.122 |
| BP medication | 0/1 | 0.00 | 0.28 | 0.45 | – | 0.156 |
| *Missingness indicators (1 = value missing)* |  |  |  |  |  |  |
| BMI, kg/m<sup>2</sup> | 0/1 | – | 0.013 | 0.113 | – | 0.051 |
| Systolic BP, mmHg | 0/1 | – | 0.043 | 0.203 | – | 0.053 |
| HDL cholesterol, mmol/l | 0/1 | – | 0.006 | 0.079 | – | 0.259 |
| Triglycerides, mmol/l | 0/1 | – | 0.006 | 0.077 | – | −0.207 |
| Current smoker | 0/1 | – | 0.003 | 0.052 | – | 0.030 |
| Weekly vigorous activity<sup>a</sup> | 0/1 | – | – | – | – | – |
| BP medication | 0/1 | – | 0.010 | 0.101 | – | 0.070 |

<sup>a</sup>Based on fewer than three baseline visits; values withheld under the UK Data Service disclosure rules. The term is included in model B and its values are available by running the analysis code on the licensed data.

**Table S5:**
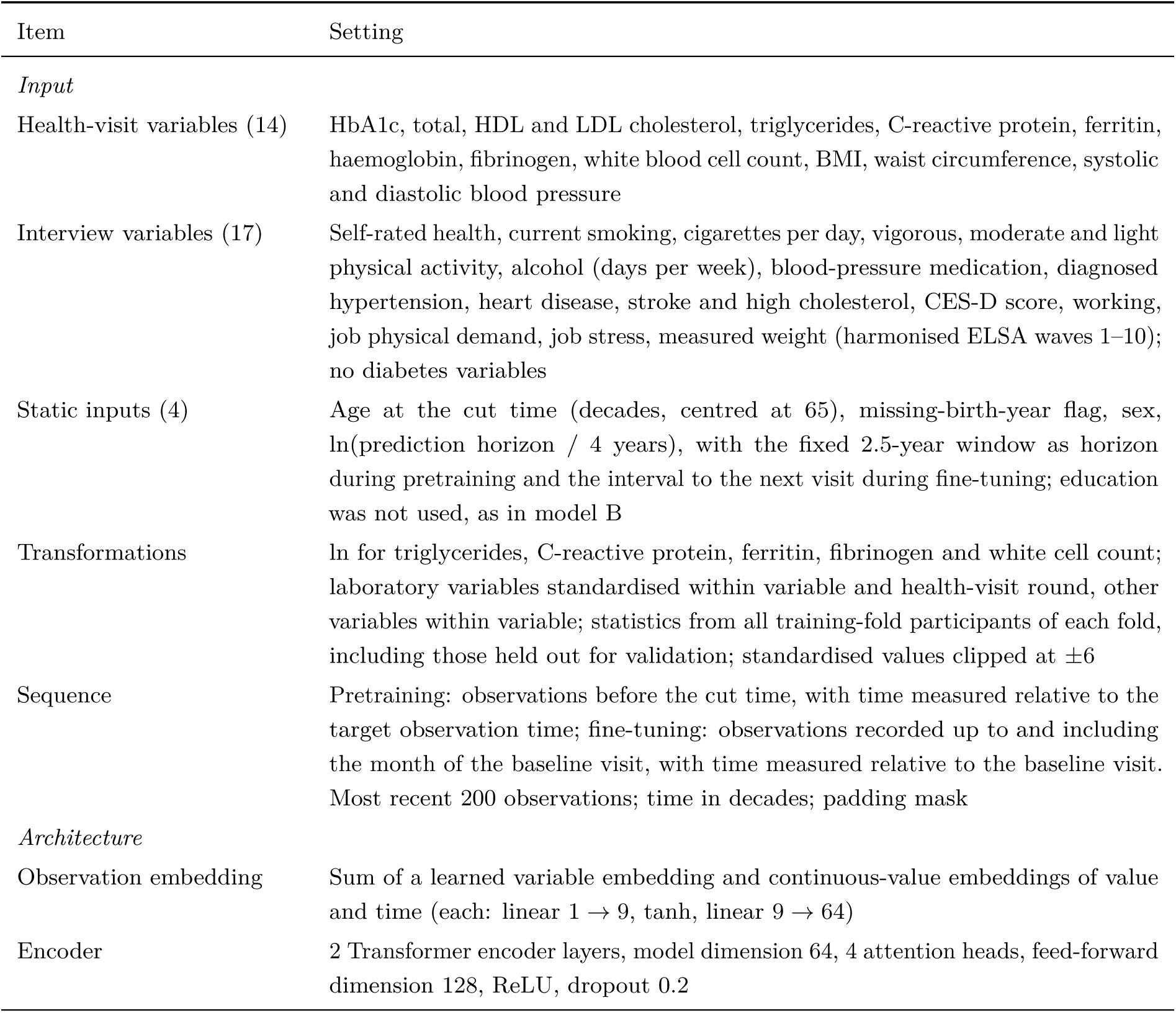
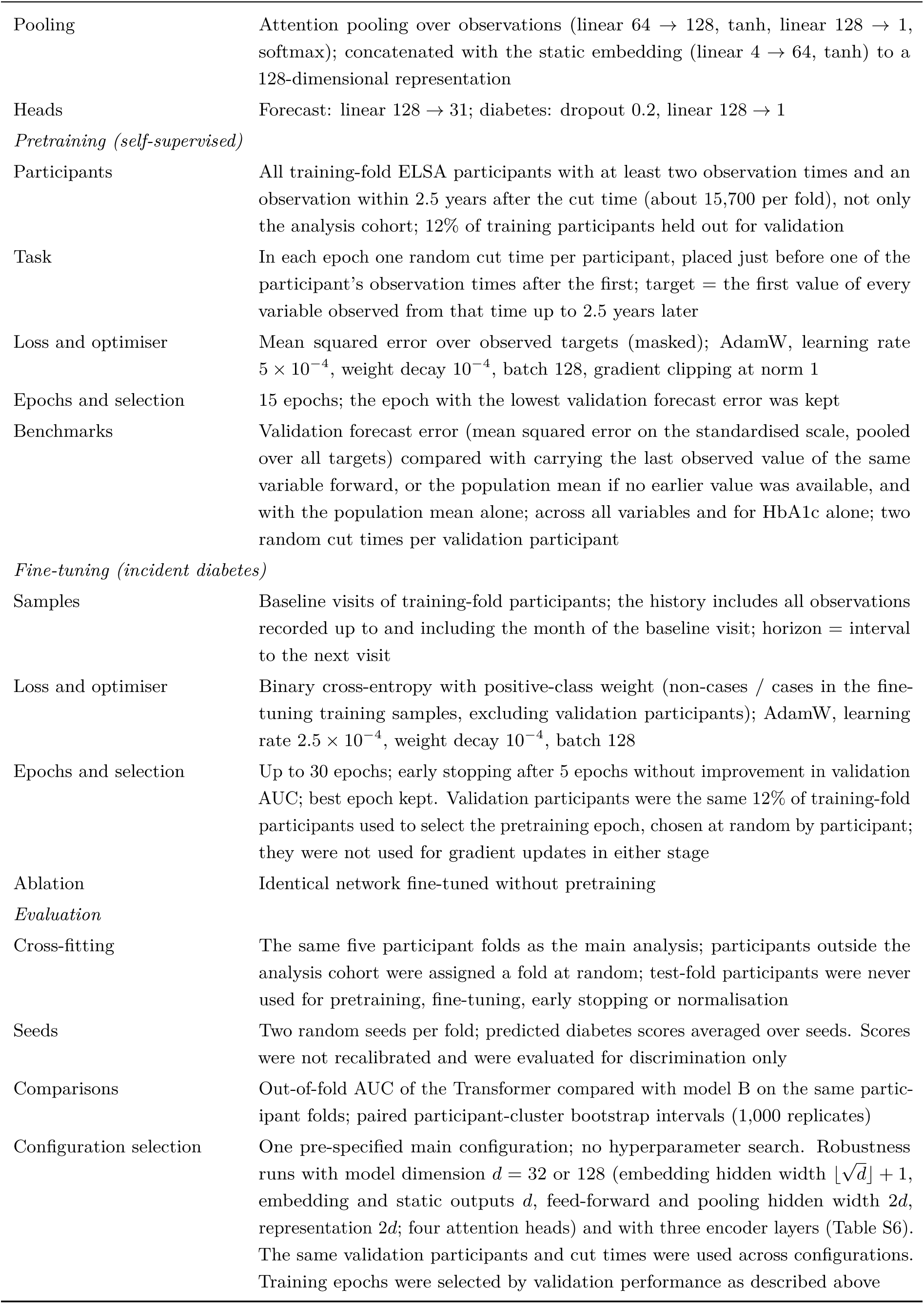
Transformer specification and training.

| Item | Setting |
|---|---|
| *Input* |  |
| Health-visit variables (14) | HbA1c, total, HDL and LDL cholesterol, triglycerides, C-reactive protein, ferritin, haemoglobin, fibrinogen, white blood cell count, BMI, waist circumference, systolic and diastolic blood pressure |
| Interview variables (17) | Self-rated health, current smoking, cigarettes per day, vigorous, moderate and light physical activity, alcohol (days per week), blood-pressure medication, diagnosed hypertension, heart disease, stroke and high cholesterol, CES-D score, working, job physical demand, job stress, measured weight (harmonised ELSA waves 1–10); no diabetes variables |
| Static inputs (4) | Age at the cut time (decades, centred at 65), missing-birth-year flag, sex, ln(prediction horizon / 4 years), with the fixed 2.5-year window as horizon during pretraining and the interval to the next visit during fine-tuning; education was not used, as in model B |
| Transformations | ln for triglycerides, C-reactive protein, ferritin, fibrinogen and white cell count; laboratory variables standardised within variable and health-visit round, other variables within variable; statistics from all training-fold participants of each fold, including those held out for validation; standardised values clipped at ±6 |
| Sequence | Pretraining: observations before the cut time, with time measured relative to the target observation time; fine-tuning: observations recorded up to and including the month of the baseline visit, with time measured relative to the baseline visit. Most recent 200 observations; time in decades; padding mask |
| *Architecture* |  |
| Observation embedding | Sum of a learned variable embedding and continuous-value embeddings of value and time (each: linear 1 → 9, tanh, linear 9 → 64) |
| Encoder | 2 Transformer encoder layers, model dimension 64, 4 attention heads, feed-forward dimension 128, ReLU, dropout 0.2 |
| Pooling | Attention pooling over observations (linear 64 → 128, tanh, linear 128 → 1, softmax); concatenated with the static embedding (linear 4 → 64, tanh) to a 128-dimensional representation |
| Heads | Forecast: linear 128 → 31; diabetes: dropout 0.2, linear 128 → 1 |
| *Pretraining (self-supervised)* |  |
| Participants | All training-fold ELSA participants with at least two observation times and an observation within 2.5 years after the cut time (about 15,700 per fold), not only the analysis cohort; 12% of training participants held out for validation |
| Task | In each epoch one random cut time per participant, placed just before one of the participant's observation times after the first; target = the first value of every variable observed from that time up to 2.5 years later |
| Loss and optimiser | Mean squared error over observed targets (masked); AdamW, learning rate 5 × 10<sup>−4</sup>, weight decay 10<sup>−4</sup>, batch 128, gradient clipping at norm 1 |
| Epochs and selection | 15 epochs; the epoch with the lowest validation forecast error was kept |
| Benchmarks | Validation forecast error (mean squared error on the standardised scale, pooled over all targets) compared with carrying the last observed value of the same variable forward, or the population mean if no earlier value was available, and with the population mean alone; across all variables and for HbA1c alone; two random cut times per validation participant |
| *Fine-tuning (incident diabetes)* |  |
| Samples | Baseline visits of training-fold participants; the history includes all observations recorded up to and including the month of the baseline visit; horizon = interval to the next visit |
| Loss and optimiser | Binary cross-entropy with positive-class weight (non-cases / cases in the fine-tuning training samples, excluding validation participants); AdamW, learning rate 2.5 × 10<sup>−4</sup>, weight decay 10<sup>−4</sup>, batch 128 |
| Epochs and selection | Up to 30 epochs; early stopping after 5 epochs without improvement in validation AUC; best epoch kept. Validation participants were the same 12% of training-fold participants used to select the pretraining epoch, chosen at random by participant; they were not used for gradient updates in either stage |
| Ablation | Identical network fine-tuned without pretraining |
| *Evaluation* |  |
| Cross-fitting | The same five participant folds as the main analysis; participants outside the analysis cohort were assigned a fold at random; test-fold participants were never used for pretraining, fine-tuning, early stopping or normalisation |
| Seeds | Two random seeds per fold; predicted diabetes scores averaged over seeds. Scores were not recalibrated and were evaluated for discrimination only |
| Comparisons | Out-of-fold AUC of the Transformer compared with model B on the same partic-ipant folds; paired participant-cluster bootstrap intervals (1,000 replicates) |
| Configuration selection | One pre-specified main configuration; no hyperparameter search. Robustness runs with model dimension *d* = 32 or 128 (embedding hidden width ⌊√*d*⌋ + 1, embedding and static outputs *d*, feed-forward and pooling hidden width 2*d*, representation 2*d*; four attention heads) and with three encoder layers (Table S6). The same validation participants and cut times were used across configurations. Training epochs were selected by validation performance as described above |

**Table S6:** Transformer robustness across alternative configurations. Out-of-fold AUCs are shown for pretrained and randomly initialised networks evaluated on 5,828 baseline visits with 234 incident cases. Differences from model B (AUC 0.837, 95% CI 0.806–0.868) refer to pretrained networks, with 95% CIs from a paired participant-cluster bootstrap. Forecast results show the percentage change in validation MSE relative to the baseline defined below, as the range across the five folds.

|  | AUC |  |  | Forecast MSE change vs baseline, % |
|---|---|---|---|---|
| Configuration | Pretrained | From scratch | ΔAUC vs model B, pretrained (95% CI) | All variables / HbA1c |
| Main (dimension 64, 2 layers) | 0.847 | 0.849 | +0.010 (−0.006 to 0.026) | −18 to −16 / −5 to +9 |
| Dimension 32 | 0.832 | 0.836 | −0.005 (−0.020 to 0.012) | −12 to −10 / +18 to +26 |
| Dimension 128 | 0.828 | 0.842 | −0.009 (−0.029 to 0.010) | −20 to −19 / −9 to +1 |
| Three layers | 0.836 | 0.841 | −0.001 (−0.021 to 0.018) | −19 to −17 / −5 to +5 |

Forecast targets were the first observed value of each variable within 2.5 years after the cut time, for two cut times per validation participant (about 3,300 samples and 63,000 targets per fold). The forecast baseline carried forward the last observed value of the same variable on its standardised scale, using the training-fold mean when no earlier value was available. Percentage changes are 100(MSE<sub>Transformer</sub>/MSE<sub>baseline</sub> − 1); negative values favour the Transformer. HbA1c contributed 1,521–1,619 targets per fold (2.4–2.6% of all targets), reflecting its measurement only at the health visits, about four years apart, whereas interview variables were recorded every two years; about half had an earlier HbA1c value. Aggregate forecast performance was therefore dominated by variables other than HbA1c, and the HbA1c comparison varied between folds.

**Table S7:**
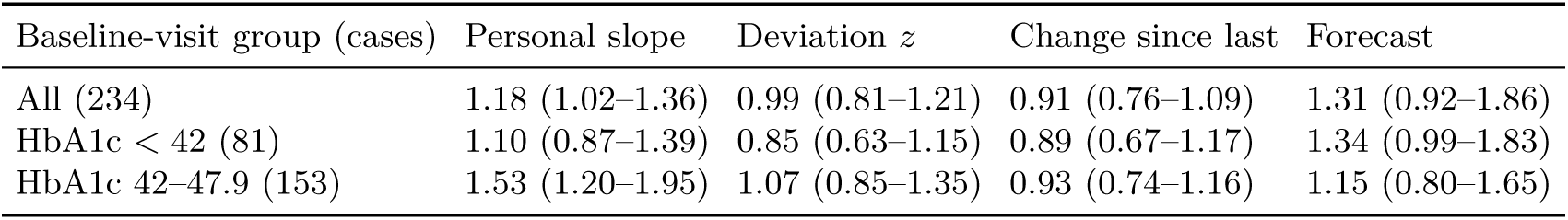
Odds ratio per SD of each personal feature (95% CI), adjusted for a spline in HbA1c, the clinical covariates of model B and ln(years to next visit). HbA1c ranges in mmol/mol. Each feature is standardised by its SD within the row’s set of baseline visits (subgroup-specific SD); estimates per SD in all baseline visits, and for baseline visits with at least two earlier values, are given in Table S8.

| Baseline-visit group (cases) | Personal slope | Deviation *z* | Change since last | Forecast |
|---|---|---|---|---|
| All (234) | 1.18 (1.02–1.36) | 0.99 (0.81–1.21) | 0.91 (0.76–1.09) | 1.31 (0.92–1.86) |
| HbA1c < 42 (81) | 1.10 (0.87–1.39) | 0.85 (0.63–1.15) | 0.89 (0.67–1.17) | 1.34 (0.99–1.83) |
| HbA1c 42–47.9 (153) | 1.53 (1.20–1.95) | 1.07 (0.85–1.35) | 0.93 (0.74–1.16) | 1.15 (0.80–1.65) |

**Table S8:**
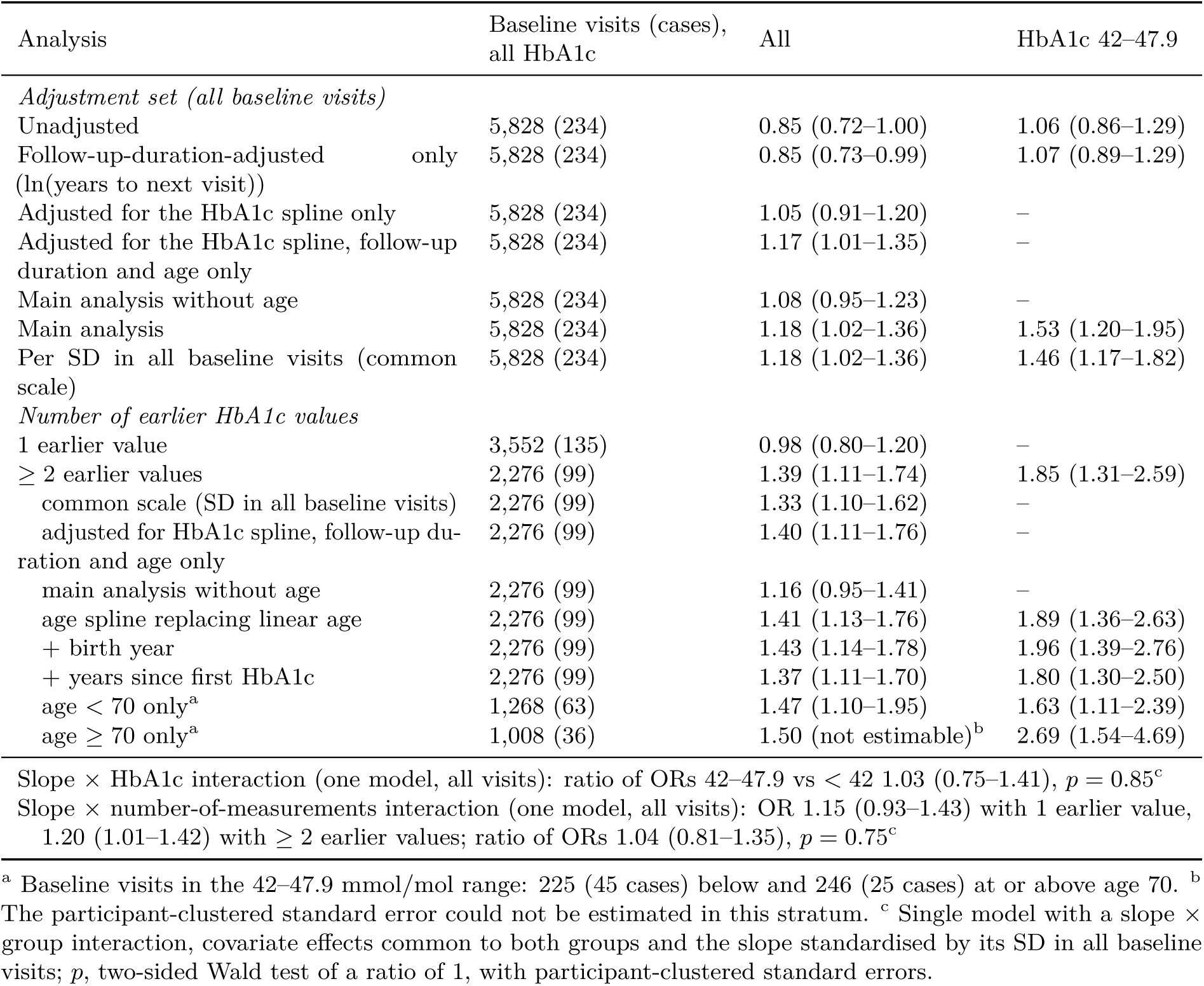
Personal-slope association: adjustment set and measurement history. Odds ratio per SD (95% CI), with the common specification described at the start of this section unless stated otherwise.

| Analysis | Baseline visits (cases), all HbA1c | All | HbA1c 42–47.9 |
|---|---|---|---|
| *Adjustment set (all baseline visits)* |  |  |  |
| Unadjusted | 5,828 (234) | 0.85 (0.72–1.00) | 1.06 (0.86–1.29) |
| Follow-up-duration-adjusted only (ln(years to next visit)) | 5,828 (234) | 0.85 (0.73–0.99) | 1.07 (0.89–1.29) |
| Adjusted for the HbA1c spline only | 5,828 (234) | 1.05 (0.91–1.20) | – |
| Adjusted for the HbA1c spline, follow-up duration and age only | 5,828 (234) | 1.17 (1.01–1.35) | – |
| Main analysis without age | 5,828 (234) | 1.08 (0.95–1.23) | – |
| Main analysis | 5,828 (234) | 1.18 (1.02–1.36) | 1.53 (1.20–1.95) |
| Per SD in all baseline visits (common scale) | 5,828 (234) | 1.18 (1.02–1.36) | 1.46 (1.17–1.82) |
| *Number of earlier HbA1c values* |  |  |  |
| 1 earlier value | 3,552 (135) | 0.98 (0.80–1.20) | – |
| ≥ 2 earlier values | 2,276 (99) | 1.39 (1.11–1.74) | 1.85 (1.31–2.59) |
| common scale (SD in all baseline visits) | 2,276 (99) | 1.33 (1.10–1.62) | – |
| adjusted for HbA1c spline, follow-up du-ration and age only | 2,276 (99) | 1.40 (1.11–1.76) | – |
| main analysis without age | 2,276 (99) | 1.16 (0.95–1.41) | – |
| age spline replacing linear age | 2,276 (99) | 1.41 (1.13–1.76) | 1.89 (1.36–2.63) |
| + birth year | 2,276 (99) | 1.43 (1.14–1.78) | 1.96 (1.39–2.76) |
| + years since first HbA1c | 2,276 (99) | 1.37 (1.11–1.70) | 1.80 (1.30–2.50) |
| age < 70 only<sup>a</sup> | 1,268 (63) | 1.47 (1.10–1.95) | 1.63 (1.11–2.39) |
| age ≥ 70 only<sup>a</sup> | 1,008 (36) | 1.50 (not estimable)<sup>b</sup> | 2.69 (1.54–4.69) |
| Slope × HbA1c interaction (one model, all visits): ratio of ORs 42–47.9 vs < 42 1.03 (0.75–1.41), *p* = 0.85<sup>c</sup> |  |  |  |
| Slope × number-of-measurements interaction (one model, all visits): OR 1.15 (0.93–1.43) with 1 earlier value, 1.20 (1.01–1.42) with ≥ 2 earlier values; ratio of ORs 1.04 (0.81–1.35), *p* = 0.75<sup>c</sup> |  |  |  |

<sup>a</sup> Baseline visits in the 42–47.9 mmol/mol range: 225 (45 cases) below and 246 (25 cases) at or above age 70. <sup>b</sup> The participant-clustered standard error could not be estimated in this stratum. <sup>c</sup> Single model with a slope × group interaction, covariate effects common to both groups and the slope standardised by its SD in all baseline visits; *p*, two-sided Wald test of a ratio of 1, with participant-clustered standard errors.

**Table S9:**
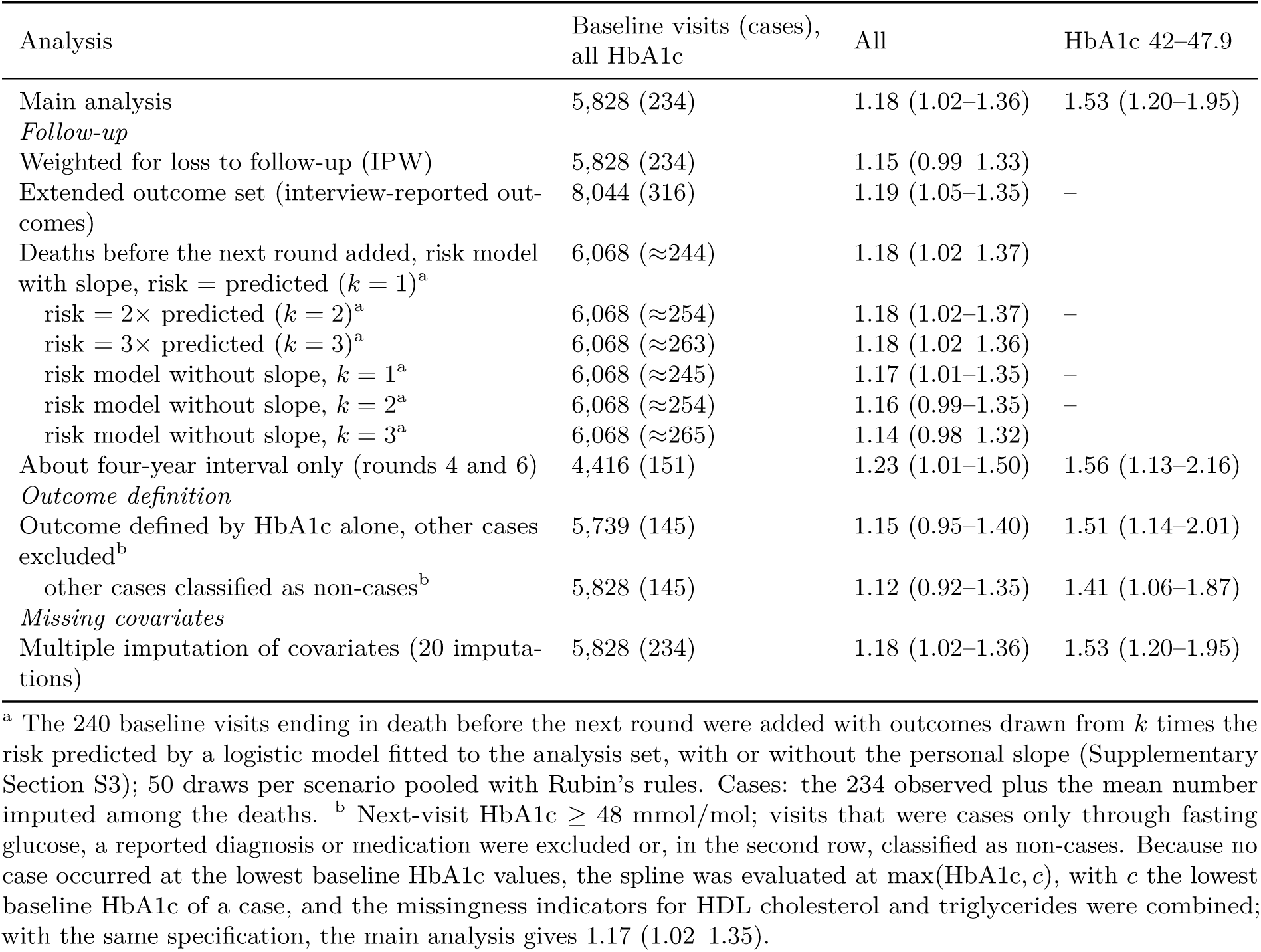
Personal-slope association: follow-up, outcome definition and missing covariates. Odds ratio per SD (95% CI), with the common specification described at the start of this section unless stated otherwise.

**Table S10:** Personal-slope association: adjustment for the current HbA1c level and construction of the slope. Odds ratio per SD (95% CI), with the common specification described at the start of this section unless stated otherwise. In the drift-model rows, the spline in baseline HbA1c is replaced by a spline in the drift model’s estimate of the underlying current level (Section 2.8).

| Analysis | Baseline visits (cases), all HbA1c | All | HbA1c 42–47.9 |
|---|---|---|---|
| *Slope from earlier values (main analysis), adjusted for* |  |  |  |
| Baseline HbA1c (main analysis) | 5,828 (234) | 1.18 (1.02–1.36) | 1.53 (1.20–1.95) |
| Mean of baseline and preceding value (corrected for round effects) | 5,828 (234) | 1.16 (1.01–1.34) | 1.56 (1.22–2.00) |
| Drift-model level, crossing values observed | 5,828 (234) | 1.20 (1.03–1.39) | 1.60 (1.24–2.08) |
| Drift-model level, crossing values censored | 5,828 (234) | 1.24 (1.07–1.44) | 1.72 (1.33–2.23) |
| *Slope including the baseline value, adjusted for* |  |  |  |
| Baseline HbA1c | 5,828 (234) | 1.10 (0.96–1.26) | 1.32 (1.08–1.61) |
| Drift-model level, crossing values observed | 5,828 (234) | 1.17 (1.02–1.35) | 1.47 (1.19–1.81) |

**Table S11:** Drift model by cap on the threshold-crossing values (non-diabetic visits plus crossing value; REML), and with the crossing values censored at 48 mmol/mol (maximum likelihood). 401 crossing values in total. CI from the profile likelihood, interpolated on log *τ* between grid values (coarser grid than in Table 4). ΔAIC = AIC(drift) AIC(independent noise) within each data set; values are not comparable between rows, which use different data. –, not computed.

| Cap,<br>mmol/mol | Crossing values<br>above cap, n (%) | τ, years (95% CI) | Slope SD,<br>mmol/mol per decade | Drift<br>share π | ΔAIC |
|---|---|---|---|---|---|
| 48 | 338 (84.3) | 29.0 (14.9–50.0) | 0.27 | 0.77 | −221 |
| 50 | 215 (53.6) | 25.0 (14.3–38.1) | 0.37 | 0.77 | −241 |
| 52 | 149 (37.2) | 21.8 (13.4–33.5) | 0.43 | 0.76 | −253 |
| 55 | 104 (25.9) | 18.4 (12.0–26.9) | 0.50 | 0.75 | −274 |
| 58 | 72 (18.0) | 16.4 (11.2–23.7) | 0.56 | 0.75 | −292 |
| 60 | 56 (14.0) | 15.0 (10.5–21.6) | 0.59 | 0.75 | −303 |
| 65 | 40 (10.0) | 13.3 (9.5–18.3) | 0.66 | 0.75 | −319 |
| 70 | 31 (7.7) | 11.7 (8.6–15.7) | 0.70 | 0.76 | −348 |
| 80 | 16 (4.0) | 9.6 (7.4–12.8) | 0.76 | 0.78 | −393 |
| None | 0 | 5.8 (4.3–7.8) | 0.81 | 0.81 | −378 |
| Censored at 48 | – | 26.0 (14.4–43.2) | 0.33 | 0.77 | −228 |

**Table S12:** Expected correlation between estimated and underlying personal slopes with the intercept– slope correlation set to zero (reference observability calculation) and at its fitted value. Equally spaced measurements, the last at age 65 except where noted. Calculations condition on the specified covariance parameters and do not incorporate their estimation uncertainty. Four-yearly schedules over 4–12 years approximate the spacing and duration of the earlier ELSA health-visit histories.

|  |  | Four-yearly values |  | Quarterly values |  |
|---|---|---|---|---|---|
| Model (fitted ρ) | History | ρ = 0 | fitted ρ | ρ = 0 | fitted ρ |
| Independent noise (−0.03) | 4 years | 0.15 | 0.17 | 0.26 | 0.27 |
|  | 12 years | 0.44 | 0.45 | 0.79 | 0.79 |
|  | 30 years<sup>b</sup> | 0.81 | 0.82 | 0.98 | 0.98 |
| Drift, with crossing values (≈ 1) | 4 years | 0.10 | 0.75 | 0.11 | 0.77 |
|  | 12 years | 0.26 | 0.78 | 0.28 | 0.80 |
|  | 30 years<sup>b</sup> | 0.56 | 0.78 | 0.59 | 0.81 |
| Drift, crossing values censored<sup>a</sup> (≈ 1) | 4 years | 0.06 | 0.69 | 0.08 | 0.71 |
|  | 12 years | 0.15 | 0.70 | 0.18 | 0.72 |
|  | 30 years<sup>b</sup> | 0.31 | 0.70 | 0.34 | 0.73 |

<sup>a</sup> Maximum-likelihood fit with the crossing values censored at 48 mmol/mol (τ = 26 years). <sup>b</sup> Four-yearly values at ages 35 to 63 (eight values, 28-year span), because four-year intervals do not divide 30 years; the quarterly schedule spans the full 30 years.

**Table S13:**
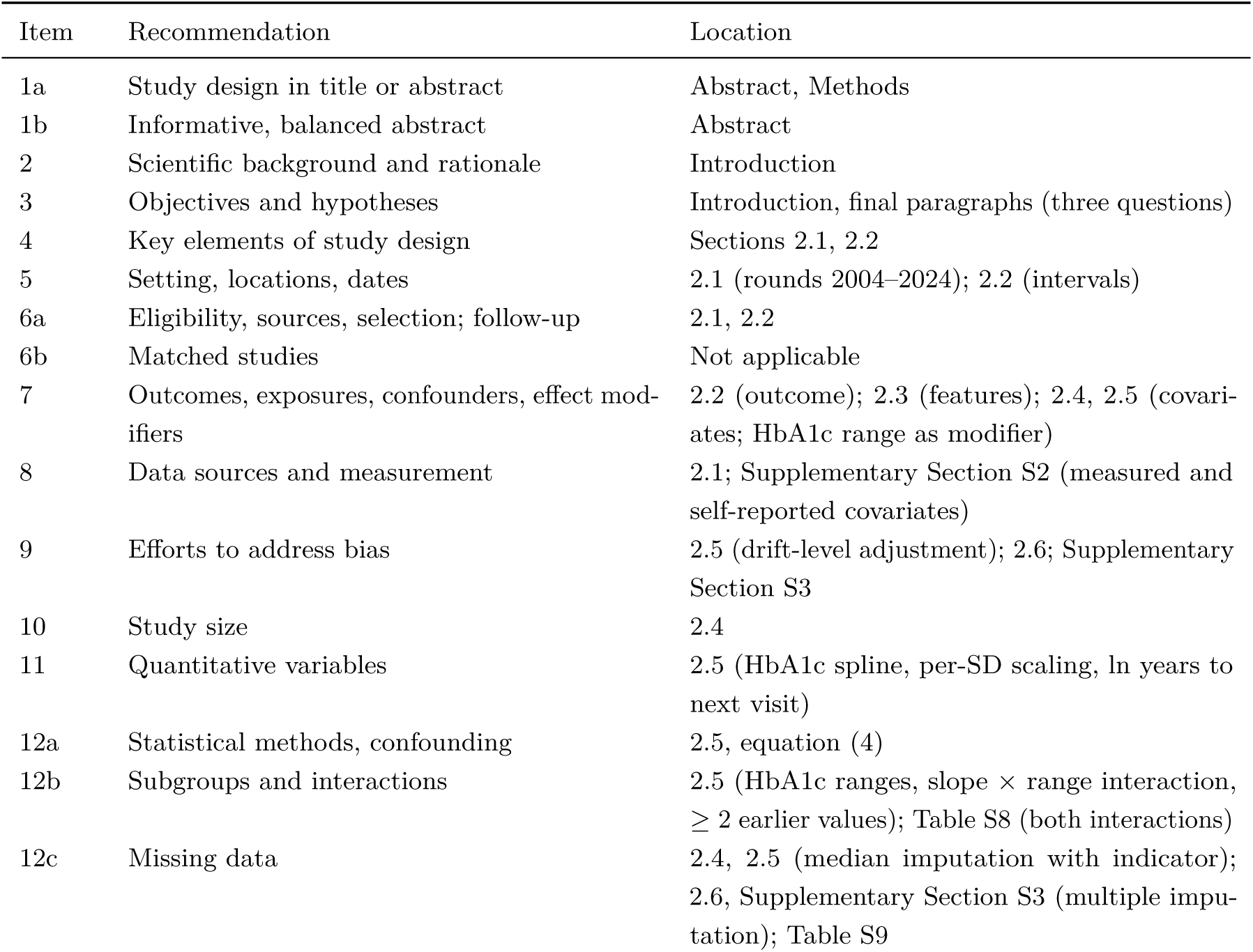
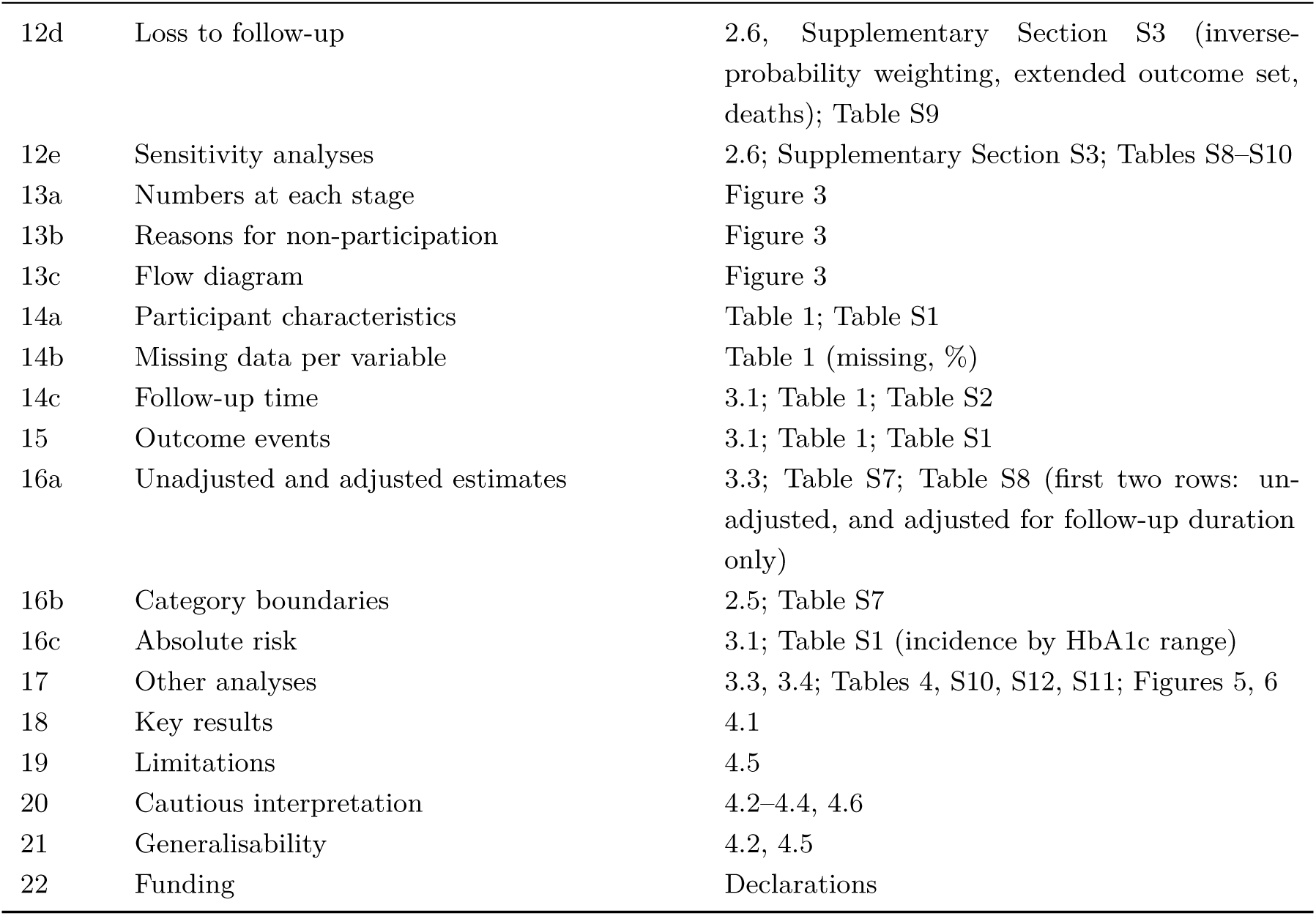
STROBE checklist for cohort studies (von Elm et al., 2007), applied to the association analyses. Item wording paraphrased.

| Item | Recommendation | Location |
|---|---|---|
| 1a | Study design in title or abstract | Abstract, Methods |
| 1b | Informative, balanced abstract | Abstract |
| 2 | Scientific background and rationale | Introduction |
| 3 | Objectives and hypotheses | Introduction, final paragraphs (three questions) |
| 4 | Key elements of study design | Sections 2.1, 2.2 |
| 5 | Setting, locations, dates | 2.1 (rounds 2004–2024); 2.2 (intervals) |
| 6a | Eligibility, sources, selection; follow-up | 2.1, 2.2 |
| 6b | Matched studies | Not applicable |
| 7 | Outcomes, exposures, confounders, effect mod-ifiers | 2.2 (outcome); 2.3 (features); 2.4, 2.5 (covari-ates; HbA1c range as modifier) |
| 8 | Data sources and measurement | 2.1; Supplementary Section S2 (measured and self-reported covariates) |
| 9 | Efforts to address bias | 2.5 (drift-level adjustment); 2.6; Supplementary Section S3 |
| 10 | Study size | 2.4 |
| 11 | Quantitative variables | 2.5 (HbA1c spline, per-SD scaling, ln years to next visit) |
| 12a | Statistical methods, confounding | 2.5, equation (4) |
| 12b | Subgroups and interactions | 2.5 (HbA1c ranges, slope × range interaction, ≥ 2 earlier values); Table S8 (both interactions) |
| 12c | Missing data | 2.4, 2.5 (median imputation with indicator); 2.6, Supplementary Section S3 (multiple impu-tation); Table S9 |
| 12d | Loss to follow-up | 2.6, Supplementary Section S3 (inverse-probability weighting, extended outcome set, deaths); Table S9 |
| 12e | Sensitivity analyses | 2.6; Supplementary Section S3; Tables S8–S10 |
| 13a | Numbers at each stage | Figure 3 |
| 13b | Reasons for non-participation | Figure 3 |
| 13c | Flow diagram | Figure 3 |
| 14a | Participant characteristics | Table 1; Table S1 |
| 14b | Missing data per variable | Table 1 (missing, %) |
| 14c | Follow-up time | 3.1; Table 1; Table S2 |
| 15 | Outcome events | 3.1; Table 1; Table S1 |
| 16a | Unadjusted and adjusted estimates | 3.3; Table S7; Table S8 (first two rows: un-adjusted, and adjusted for follow-up duration only) |
| 16b | Category boundaries | 2.5; Table S7 |
| 16c | Absolute risk | 3.1; Table S1 (incidence by HbA1c range) |
| 17 | Other analyses | 3.3, 3.4; Tables 4, S10, S12, S11; Figures 5, 6 |
| 18 | Key results | 4.1 |
| 19 | Limitations | 4.5 |
| 20 | Cautious interpretation | 4.2–4.4, 4.6 |
| 21 | Generalisability | 4.2, 4.5 |
| 22 | Funding | Declarations |

**Table S14:**
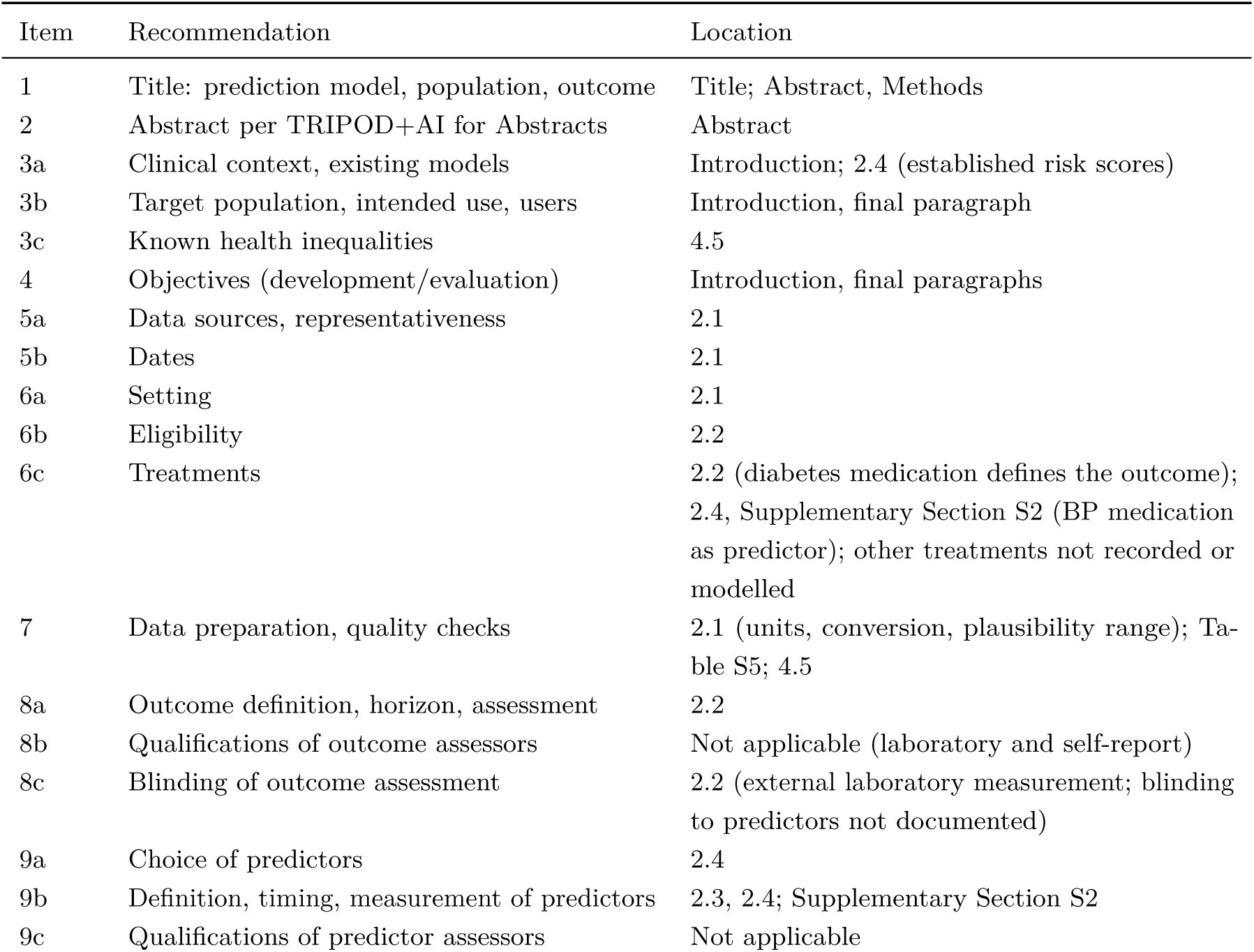
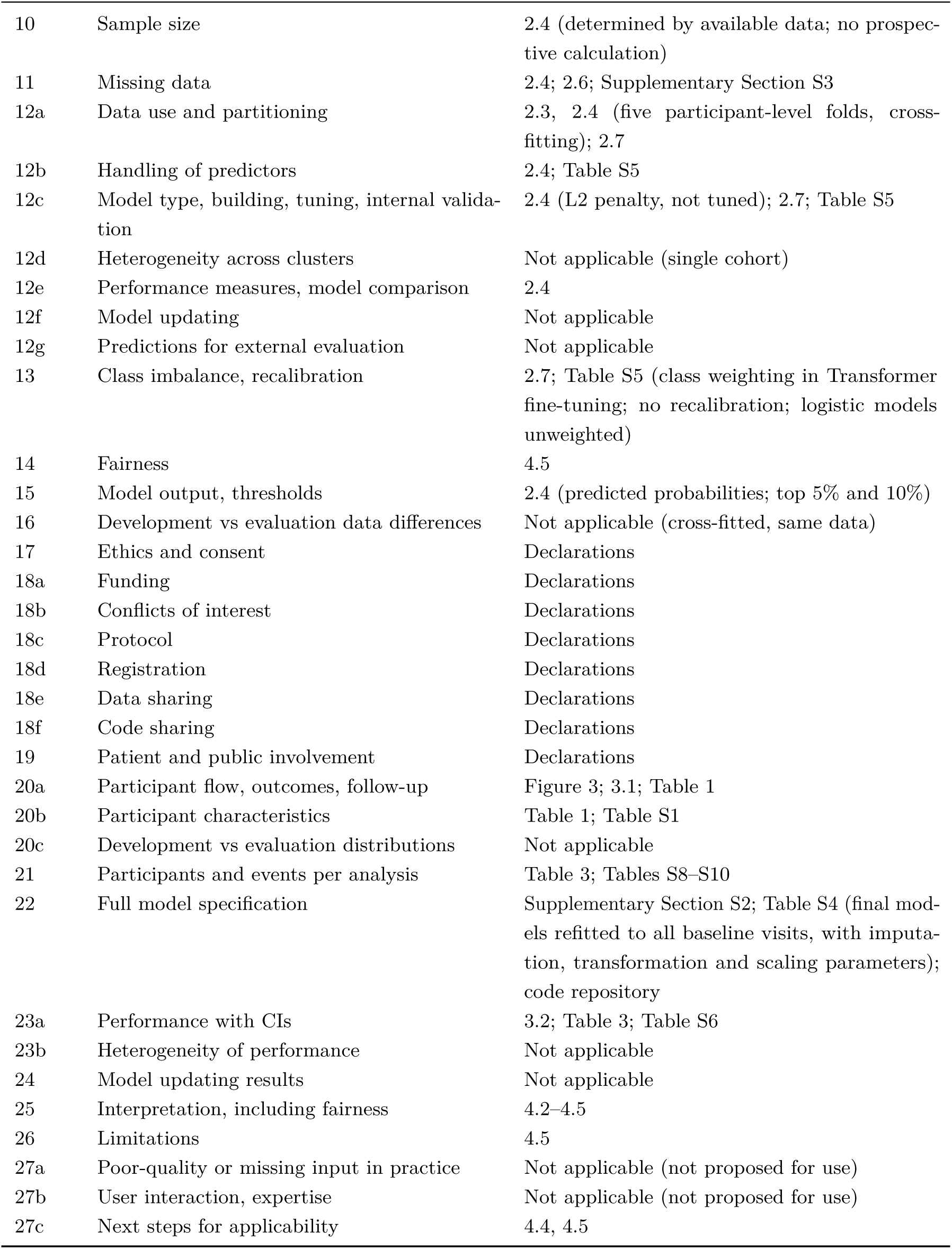
TRIPOD+AI checklist (Collins et al., 2024), applied to the prediction analyses. Item wording paraphrased. Items on external evaluation do not apply: all performance is internal and out of fold.

| Item | Recommendation | Location |
|---|---|---|
| 1 | Title: prediction model, population, outcome | Title; Abstract, Methods |
| 2 | Abstract per TRIPOD+AI for Abstracts | Abstract |
| 3a | Clinical context, existing models | Introduction; 2.4 (established risk scores) |
| 3b | Target population, intended use, users | Introduction, final paragraph |
| 3c | Known health inequalities | 4.5 |
| 4 | Objectives (development/evaluation) | Introduction, final paragraphs |
| 5a | Data sources, representativeness | 2.1 |
| 5b | Dates | 2.1 |
| 6a | Setting | 2.1 |
| 6b | Eligibility | 2.2 |
| 6c | Treatments | 2.2 (diabetes medication defines the outcome); 2.4, Supplementary Section S2 (BP medication as predictor); other treatments not recorded or modelled |
| 7 | Data preparation, quality checks | 2.1 (units, conversion, plausibility range); Ta-ble S5; 4.5 |
| 8a | Outcome definition, horizon, assessment | 2.2 |
| 8b | Qualifications of outcome assessors | Not applicable (laboratory and self-report) |
| 8c | Blinding of outcome assessment | 2.2 (external laboratory measurement; blinding to predictors not documented) |
| 9a | Choice of predictors | 2.4 |
| 9b | Definition, timing, measurement of predictors | 2.3, 2.4; Supplementary Section S2 |
| 9c | Qualifications of predictor assessors | Not applicable |
| 10 | Sample size | 2.4 (determined by available data; no prospec-tive calculation) |
| 11 | Missing data | 2.4; 2.6; Supplementary Section S3 |
| 12a | Data use and partitioning | 2.3, 2.4 (five participant-level folds, cross-fitting); 2.7 |
| 12b | Handling of predictors | 2.4; Table S5 |
| 12c | Model type, building, tuning, internal valida-tion | 2.4 (L2 penalty, not tuned); 2.7; Table S5 |
| 12d | Heterogeneity across clusters | Not applicable (single cohort) |
| 12e | Performance measures, model comparison | 2.4 |
| 12f | Model updating | Not applicable |
| 12g | Predictions for external evaluation | Not applicable |
| 13 | Class imbalance, recalibration | 2.7; Table S5 (class weighting in Transformer fine-tuning; no recalibration; logistic models unweighted) |
| 14 | Fairness | 4.5 |
| 15 | Model output, thresholds | 2.4 (predicted probabilities; top 5% and 10%) |
| 16 | Development vs evaluation data differences | Not applicable (cross-fitted, same data) |
| 17 | Ethics and consent | Declarations |
| 18a | Funding | Declarations |
| 18b | Conflicts of interest | Declarations |
| 18c | Protocol | Declarations |
| 18d | Registration | Declarations |
| 18e | Data sharing | Declarations |
| 18f | Code sharing | Declarations |
| 19 | Patient and public involvement | Declarations |
| 20a | Participant flow, outcomes, follow-up | Figure 3; 3.1; Table 1 |
| 20b | Participant characteristics | Table 1; Table S1 |
| 20c | Development vs evaluation distributions | Not applicable |
| 21 | Participants and events per analysis | Table 3; Tables S8–S10 |
| 22 | Full model specification | Supplementary Section S2; Table S4 (final mod-els refitted to all baseline visits, with imputa-tion, transformation and scaling parameters); code repository |
| 23a | Performance with CIs | 3.2; Table 3; Table S6 |
| 23b | Heterogeneity of performance | Not applicable |
| 24 | Model updating results | Not applicable |
| 25 | Interpretation, including fairness | 4.2–4.5 |
| 26 | Limitations | 4.5 |
| 27a | Poor-quality or missing input in practice | Not applicable (not proposed for use) |
| 27b | User interaction, expertise | Not applicable (not proposed for use) |
| 27c | Next steps for applicability | 4.4, 4.5 |

## Footnotes

1 Other guidelines use a lower bound of 39 mmol/mol (5.7%) (American Diabetes Association Professional Practice Committee, 2026; Landgraf et al., 2025).

